# Leveraging large-scale biobank EHRs to enhance pharmacogenetics of cardiometabolic disease medications

**DOI:** 10.1101/2024.04.06.24305415

**Authors:** Marie C. Sadler, Alexander Apostolov, Caterina Cevallos, Diogo M. Ribeiro, Russ B. Altman, Zoltán Kutalik

**Affiliations:** University Center for Primary Care and Public Health, Lausanne, Switzerland; Swiss Institute of Bioinformatics, Lausanne, Switzerland; Department of Computational Biology, University of Lausanne, Lausanne, Switzerland; Center for Integrative Genomics, University of Lausanne, Lausanne, Switzerland; Department of Bioengineering, Stanford University, Stanford, CA, USA

## Abstract

Electronic health records (EHRs) coupled with large-scale biobanks offer great promises to unravel the genetic underpinnings of treatment efficacy. However, medication-induced biomarker trajectories stemming from such records remain poorly studied. Here, we extract clinical and medication prescription data from EHRs and conduct GWAS and rare variant burden tests in the UK Biobank (discovery) and the All of Us program (replication) on ten cardiometabolic drug response outcomes including lipid response to statins, HbA1c response to metformin and blood pressure response to antihypertensives (N = 740-26,669). Our findings at genome-wide significance level recover previously reported pharmacogenetic signals and also include novel associations for lipid response to statins (N = 26,669) near *LDLR* and *ZNF800*. Importantly, these associations are treatment-specific and not associated with biomarker progression in medication-naive individuals. Furthermore, we demonstrate that individuals with higher genetically determined low-density and total cholesterol baseline levels experience increased absolute, albeit lower relative biomarker reduction following statin treatment. In summary, we systematically investigated the common and rare pharmacogenetic contribution to cardiometabolic drug response phenotypes in over 50,000 UK Biobank and All of Us participants with EHR and identified clinically relevant genetic predictors for improved personalized treatment strategies.

## Introduction

Genetic factors can contribute to inter-individual variability in drug response. However, despite the immense progress of genome-wide association studies (GWAS) for complex traits and diseases, the scale of pharmacogenetics (PGx) studies to find genetic predictors of drug efficacy remains limited. PGx GWAS represent less than 10% of all entries in the GWAS Catalog with median sample sizes of 1,220 for PGx GWAS published between 2016 and 2020 [1]. As a result of low sample size and lack of cohorts suitable for pharmacogenomic studies, relatively few PGx associations determining drug efficacy have been identified in a genome-wide approach [1, 2, 3].

Several PGx GWAS consortia have formed over the years to study the genetics of drug efficacy in larger sample sizes. For instance, the Genomic Investigation of Statin Therapy (GIST) consortium has identified variants in the *LPA*, *APOE*, *SORT1*/*CELSR2*/*PSRC1* and *SLCO1B1* regions as modulators of low-density lipoprotein cholesterol (LDL-C) response to statins by combining randomized controlled trials (RCTs) and observational studies [4]. Similarly, the Metformin Genetics (MetGen) consortium has identified *SLC2A2* as influencing haemoglobin A1c (HbA1c) response to metformin [5], and more recently a meta-GWAS on HbA1c response to GLP-1 receptor agonists found variants in *ARRB1* to influence drug efficacy [6]. Furthermore, the International Consortium for Antihypertensive Pharmacogenomics Studies (ICAPS) has published multiple GWAS investigating blood pressure response to several antihypertensive drug classes (beta blockers, calcium channel blockers (CCBs), thiazide/thiazide-like diuretics and ACE-inhibitors (ACEi)/angiotensin receptor blockers (ARB)) [7, 8, 9].

Biobanks coupled with electronic health records (EHRs) that comprise medication data provide new opportunities to discover PGx associations [10, 1, 11]. These massive datasets have already contributed to the replication of known PGx interactions as well as the discovery of new putative associations in national biobanks such as the Estonian [12] and UK Biobank (UKBB) [13, 14]. More recently, GWAS on longitudinal medication patterns extracted from the Finnish nationwide drug purchase registry in the FinnGen study identified tens of cardiometabolic risk loci specific to medication use and not associated with the underlying indication [15]. Yet, PGx biobank studies so far have either focused on known pharmacogenes and their associations with adverse drug reactions, drug dosage and drug prescribing behaviour or analyzed the genetics of temporal medication use in isolation of disease phenotypes. What remains largely unexplored is the integration of longitudinal medication and phenotypic data to screen for genetic determinants of drug efficacy at a biobank scale.

Here, we extracted clinical and medication prescription data from EHRs and conducted PGx association analyses on the change in biomarker levels following drug therapy to treat cardiometabolic diseases (Figure 1a). We assessed associations with both common and rare variants by performing GWAS and rare variant burden tests on sequencing data. Discovery analyses were conducted in the UK Biobank (UKBB) [16] and replication analyses in the All of Us (AoU) research program [17] (Figure 1b). In follow-up analyses, we compared drug response genetics to the genetics of baseline and longitudinal biomarker changes in medication-naive individuals to dissect medication- and disease-specific components while also highlighting common pitfalls in the analysis of longitudinal (response) phenotypes (Figure 1c). Finally, we demonstrated that polygenic risk scores (PRS) of the underlying condition can predict drug response. In summary, we present a comprehensive resource on the genetic architecture of cardiometabolic drug response, introduce a more reliable model for studying genetic associations with drug response, and showcase the value of analyzing EHR-coupled biobanks with longitudinal data to study inter-individual variability in drug response.

**Figure 1:**
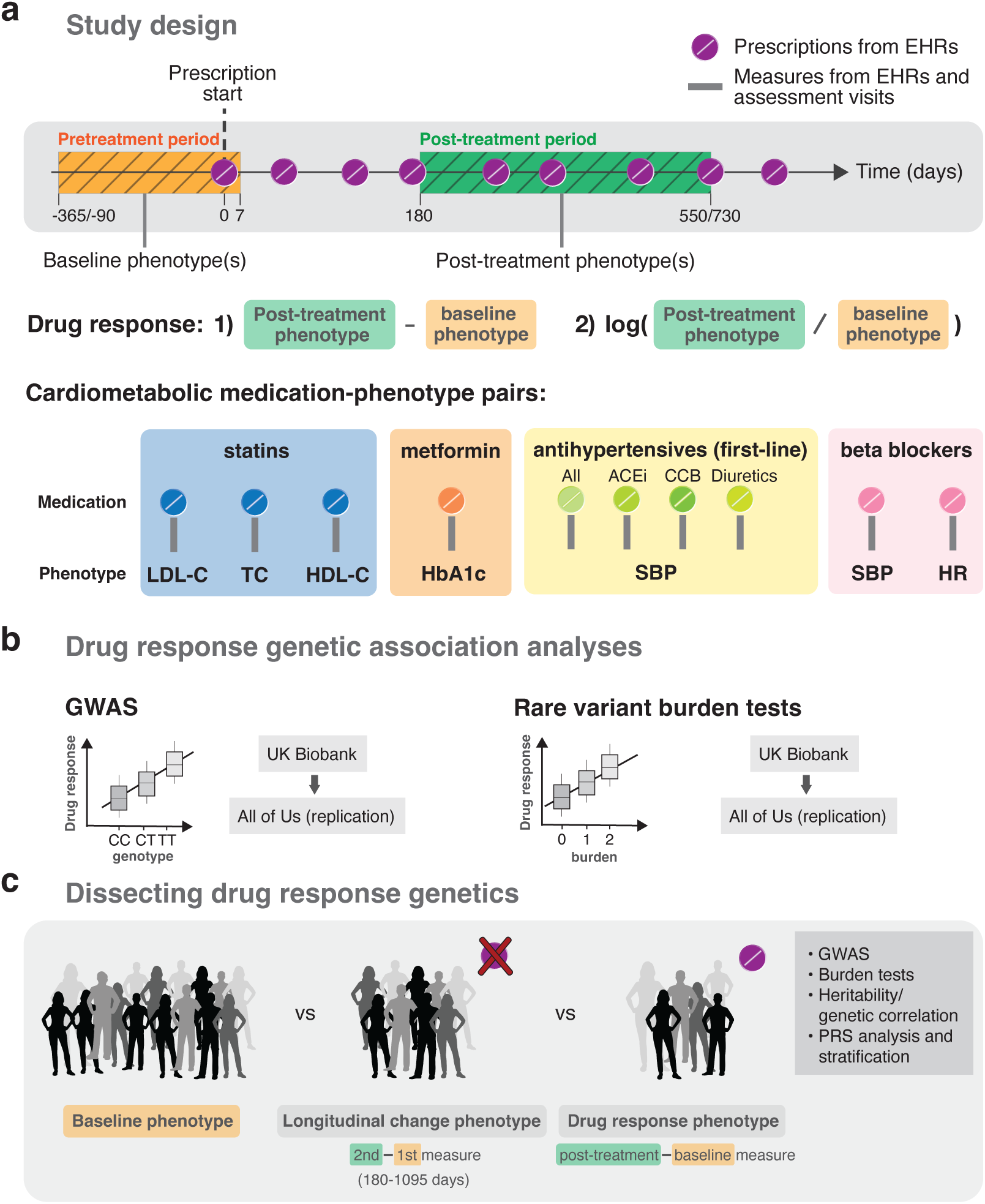
Study design. **a** Drug response study design using electronic health records (EHRs) from the UK and All of Us biobanks. Baseline and post-treatment phenotypes were extracted from EHRs or biobank assessment visits before and after the first recorded prescription, respectively. Different timings relative to the first prescription were tested as well as the use of single and average values over multiple baseline and post-treatment measures if available. Drug response phenotypes defined by the 1) absolute and 2) relative logarithmic difference in post-treatment and baseline biomarker measures were tested for ten cardiometabolic medication-phenotype pairs. **b** Discovery genetic association analyses were conducted in the UK Biobank and replicated in the All of Us research program on common variants (GWAS analysis) and rare variants through burden tests. **c** Follow-up analyses compared the genetics of baseline, longitudinal change and drug response genetics.

## Results

### Overview of the analysis

In the drug response discovery analyses, we extracted longitudinal prescription and response biomarker data from the UKBB primary care records which we combined with phenotypic data from the assessment visits. We then emulated EHR-derived drug response cohorts for the following medication-biomarker pairs: statin-lipids (LDL-C, high-density lipoprotein cholesterol (HDL-C), total cholesterol (TC)), metformin-HbA1c, antihypertensive-systolic blood pressure (SBP; by antihypertensive class (ACEi, CCB, thiazide diuretics) and all classes combined), beta blocker-SBP and beta blocker-heart rate (HR). Individuals were only part of a drug response cohort if a phenotype measurement was available before and after treatment initiation in addition to passing several other quality control (QC) steps (Method section: Study design and phenotype definitions, Figure S1, Table S3). For each drug response phenotype, we derived an absolute and logarithmic relative biomarker difference as outcome traits. Furthermore, we considered two filtering scenarios, a stringent and a lenient one. More stringent QC should result in a cleaner phenotype definition, with the trade-off of reduced sample size (and thus potentially lower statistical power). Given the sharp drop in sample size with more stringent criteria, the lenient filtering strategy constitutes the default setting throughout this study. In both stringent and lenient scenarios, we tested single and average baseline and post-treatment values over multiple measures, if available, with average values being the default (Figure 1).

In each drug response cohort, we first conducted GWAS to discover common genetic predictors (minor allele frequency (MAF) *≥* 0.05) of drug efficacy. In a second step, we performed genome-wide burden tests using whole exome sequencing (WES) data to assess associations with rare variants (MAF *<* 0.01). Replication analyses of identified PGx variants in the discovery analyses and across the literature were conducted in ∼250,000 participants of the AoU research program with available whole genome sequencing data (WGS). We also showcased the biases emerging from the popular approach of regression-based baseline adjustment to derive drug response outcomes. Finally, we assessed base-line trait PRS as predictors of drug response.

### Drug response GWAS using EHRs from the UKBB

Following cardiometabolic drug treatment, biomarker levels significantly dropped except for HDL-C which moderately increased after statin initiation (ΔHDL-C = 0.014 mmol/L, two-sided paired t-test p-value = 1.49e-24; Figure 2a; Table S4). In comparison, biomarker levels measured during an equivalent time window as baseline and post-treatment levels remained stable for control individuals who did not take any related medications (Figure 2a; Table S5). In the LDL-C response to statin GWAS, *LDLR* was found to influence absolute biomarker change (rs6511720 G*>*T, beta = -0.103, p-value = 1.29e-09), while the *SLC22A3*/*LPA* (rs10455872 A*>*G, beta = -0.148, p-value = 5.07e-16) locus and *APOE* loci (rs7412 C*>*T, beta = 0.254, p-value = 2.08e-31) were found to influence relative (logarithmic) biomarker change (Table 1; Figure 2b; lenient filtering with average values if available, N = 17,063). TC response to statins, for which we had a larger sample size (more TC than LDL-C measures are available in the primary care data, N = 26,669) confirmed the identified loci at *LDLR*, *SLC22A3*/*LPA*, and *APOE*, while also identifying a significant signal nearby *ZNF800* (rs7803925 C*>*T, beta = -0.078, p-value = 4.09e-08) for TC relative change. The SNP rs4149056 T*>*C in the *SLCO1B1* locus, also known as Val174Ala or SLCO1B1*5, which has previously been associated with LDL-C statin response [18] as well as clinical myopathy [19] barely missed genome-wide significance level (beta = -0.063, p-value = 5.33e-08). No genome-wide significant hits were found in the HDL-C response to statin GWAS (N = 23,306), HbA1c response to metformin GWAS (N = 4,119), HR (N = 1,784)/SBP (N = 1,454) response to beta blockers, and SBP response to antihypertensives (N = 740-6,933; Figures S4-5).

**Figure 2:**
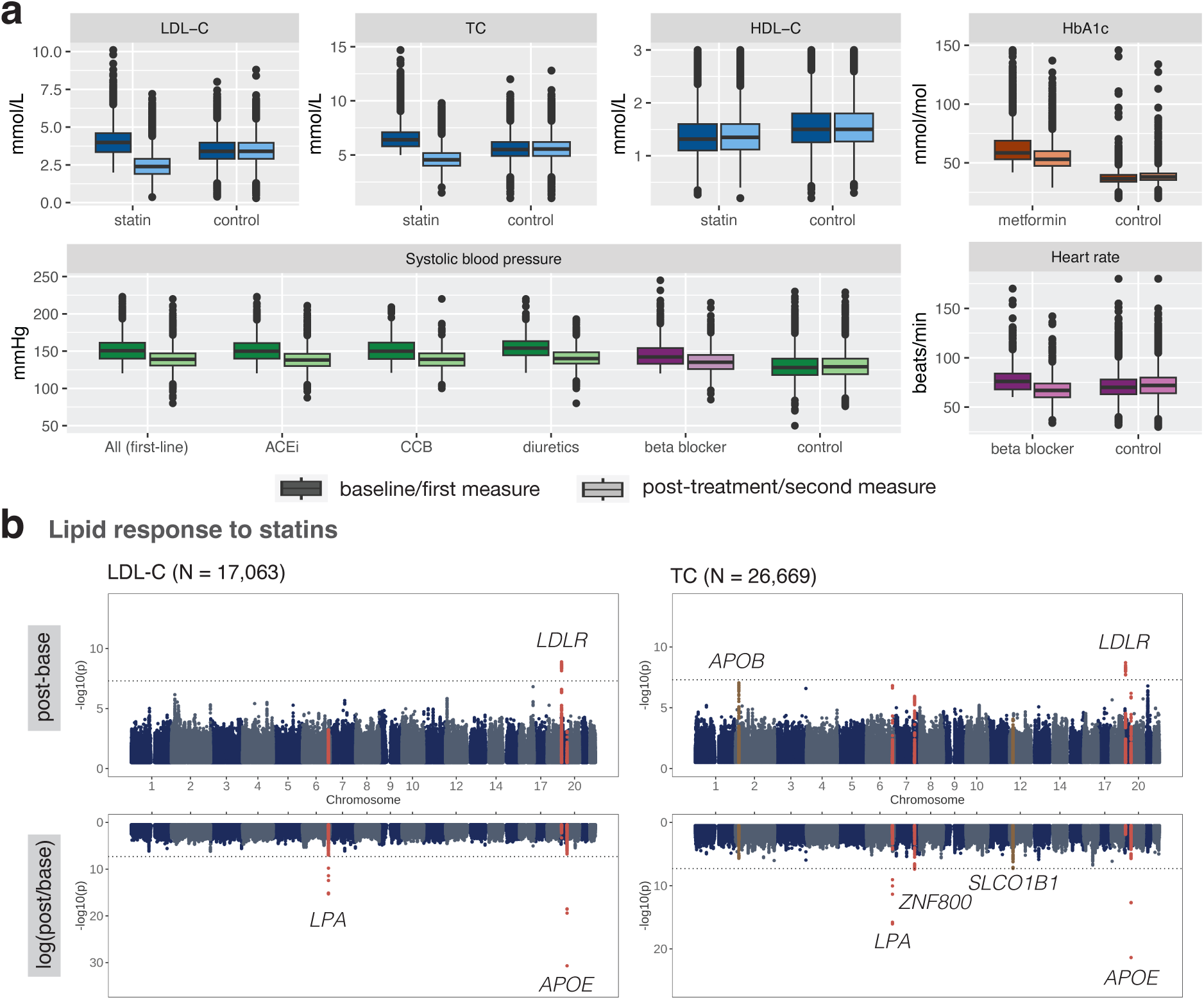
EHR drug response phenotypes and PGx GWAS results derived from the UKBB. **a** Base-line and post-treatment biomarker levels of statin (blue), metformin (orange), first-line antihypertensives (green) and beta blocker (purple) medication users as well as first and second measures of controls who do not take any related medications. **b** Manhattan plots of LDL-C and TC response to statins. GWAS association results of the top and bottom show the absolute and logarithmic relative biomarker differences, respectively. Loci with genome-wide significant signals (p-value *<* 5e-8) for either the absolute or relative difference are highlighted in red. Loci with genome-wide significant signals in other settings and with p-values *<* 1e-7 are highlighted in brown. All loci are annotated with the closest gene and the horizontal line denotes genome-wide significance (p-value *<* 5e-8). Results in **a** and **b** correspond to the lenient filtering setting with average values over multiple measures, if available.

**Table 1:** Genome-wide significant loci in discovery analyses (UK Biobank) across all assessed cardiometabolic drug response traits together with replication results (All of Us). Chr, chromosome; EAF, frequency of effect allele; post-base, absolute biomarker difference; log(post/base), logarithmic (relative) biomarker difference.

| Pharmacogenetics trait | Response definition | Chr | Position (GRCh37) | Lead SNP | Gene | Effect allele | Other allele | EAF (discovery) | N (discovery) | beta (discovery) | p-value (discovery) | EAF (replication) | N (replication) | beta (replication) | p-value (replication) |
| --- | --- | --- | --- | --- | --- | --- | --- | --- | --- | --- | --- | --- | --- | --- | --- |
| LDL-C response to statin | post-base | 19 | 11202306 | rs6511720 | LDLR | G | T | 0.894 | 17063 | -0.103 | 1.29E-09 | 0.893 | 9944 | 0.023 | 0.30 |
| LDL-C response to statin | log(post/base) | 6 | 161010118 | rs10455872 | SLC22A3, LPA | A | G | 0.910 | 17063 | -0.148 | 5.07E-16 | 0.950 | 9943 | -0.078 | 9.35E-03 |
| LDL-C response to statin | log(post/base) | 19 | 45412079 | rs7412 | APOE | C | T | 0.940 | 17063 | 0.254 | 2.08E-31 | 0.938 | 9944 | 0.205 | 4.01E-14 |
| TC response to statin | post-base | 19 | 11190481 | rs77265569 | LDLR | G | T | 0.901 | 26669 | -0.085 | 1.95E-09 | 0.887 | 6565 | -0.002 | 0.95 |
| TC response to statin | log(post/base) | 6 | 160985526 | rs118039278 | SLC22A3, LPA | G | A | 0.910 | 26669 | -0.122 | 9.08E-17 | 0.948 | 6713 | -0.070 | 4.98E-02 |
| TC response to statin | log(post/base) | 7 | 127101187 | rs7803925 | ZNF800 | C | T | 0.905 | 26669 | -0.078 | 4.09E-08 | 0.918 | 6713 | -0.019 | 0.49 |
| TC response to statin | log(post/base) | 19 | 45412079 | rs7412 | APOE | C | T | 0.941 | 26669 | 0.169 | 4.30E-22 | 0.942 | 6713 | 0.129 | 1.15E-04 |

The impact of single *vs* average baseline/post-treatment measures was minimal, and a difference was only observed for TC response to statin, with *ZNF800* reaching genome-wide significance only with average values (Figure S6-7; Table S6). The difference between stringent and lenient filtering was more pronounced, as sample sizes almost doubled with more lenient settings. For statins, this rise was largely due to the extended baseline period. For metformin and antihypertensives, we excluded individuals taking any related medication in the stringent filtering setting, whereas, in the lenient setting, sample size largely increased by allowing metformin and antihypertensives to act as add-on therapy to sulfonylureas and second-line antihypertensives, respectively, if consistently taken during pre- and post-treatment periods of the studied medication (Figures S2-3). As a consequence of lower statistical power, only 4 out of the 7 signals found in the lipid-statin GWAS were detected in the stringent filtering scenarios (Figures S6-7; Table S6). On the other hand, *APOB* was found to be significant (rs6544366 G*>*T, beta = -0.069, p-value = 3.22e-9) in the stringent filtering setting of TC response to statins while missing the genome-wide significance threshold in the lenient filtering setting (p-value = 8.90e-08).

### Replication analysis in the All of Us research program

We conducted replication analyses in the AoU program (v7; N *≈* 250,000 with available short-read WGS data). As in the UKBB, longitudinal prescription and phenotypic data were extracted from EHRs and used to construct drug response cohorts by following the same methodology as in the UKBB (Methods; Table S7). Cohort characteristics were similar to those in the UKBB (Table S7; Figure S8). The mean statin starting age was 58 years compared to 61 years in the UKBB and as in the UKBB post-treatment lipid levels were on average measured a year after the first prescription. The main difference was observed in the regularity of statin prescriptions. Whereas in the UKBB, participants had on average a prescription every two months 88% of the time, this number dropped to 42% in the AoU. There were slightly fewer statin users than in the UKBB, but similar to the UKBB, the main reasons for being excluded in the PGx cohort were missing baseline and/or post-treatment measures in the considered time windows leaving 9,944 and 6,713 individuals in the LDL-C and TC response to statins, respectively. Among the 7 signals, 2 replicated at the Bonferroni-corrected replication threshold of 0.05/7 = 0.00714 and 4 at a nominal p-value of 0.05 (all directionally concordant; Table 1). Signals not replicating nominally include the *LDLR* locus in the LDL-C and TC-response to statin GWAS as well as the *ZNF800* locus in the TC response to statin GWAS.

### EHR-derived PGx GWAS recover known PGx loci

From the literature, we extracted genetic predictors reported for the assessed cardiometabolic medication-biomarker pairs. We adopted the criteria from Nelson *et al.*, 2016 [2] that provide a curated list up to July 2015 by querying the GWAS Catalog [20]. Briefly, genetic variants were required to pass the genome-wide significance threshold of 5e-8 and show evidence of replication. Reported GWAS stem either from randomized controlled trials or observational studies often meta-analyzed together.

As we will elaborate in the longitudinal phenotype model later on (Figure 3a), adjusting biomarker changes for baseline levels induces spurious associations for genetic variants that are also associated with baseline levels. However, 4 of the 6 studies reporting significant PGx variants have adjusted for baseline levels [4, 21, 22, 23]. Thus, these reported loci could represent either baseline genetic or pharmacogenetic effects. To be concordant with these studies, we report literature replication p-values for baseline-adjusted and unadjusted biomarker change. Five independent loci were reported for LDL-C response to statins of which three (*APOE*, *LPA*, and *SORT1*) and two (*APOE* and *SORT1*) passed genome-wide significance in the (baseline adjusted) discovery (UKBB) and replication (AoU) cohort, respectively (Table 2). *SLCO1B1* locus was nominally significant in the UKBB (p-value = 8.89e-03) and genome-wide significant in the baseline adjusted TC response GWAS for which sample size was larger (p-value = 2.17e-08). *ABCG2* associated with LDL-C reduction following rosuvastatin therapy in the JUPITER trial [24] was found to be insignificant in the UKBB and AoU (p-values of *>* 0.05) and did also not reach genome-wide significance in a later, larger GWAS meta-analysis of all statins combined [4]. HDL-C response GWAS to statins (baseline adjusted) identified *CETP* as a single genome-wide significant locus, which replicated at a genome-wide significance level in the UKBB [21]. Overall, EHR-derived PGx signals on lipids agree well with those reported in cohort studies, although, baseline adjustment can lead to spurious associations for variants that associate with the baseline levels of the biomarker. For instance, rs247616 (*CETP*) was strongly associated with HDL-C change when adjusting for baseline in the UKBB (p-value = 7.77e-10), but no longer in the unadjusted analysis (p-value *>* 0.50). Similar associations of these spurious hits found in our negative control analysis (involving longitudinal change in drug-naive participants; Figure S13) confirms our suspicion that baseline adjustment leads to biased results (see Section on *Modelling drug response and longitudinal change phenotypes*).

**Figure 3:**
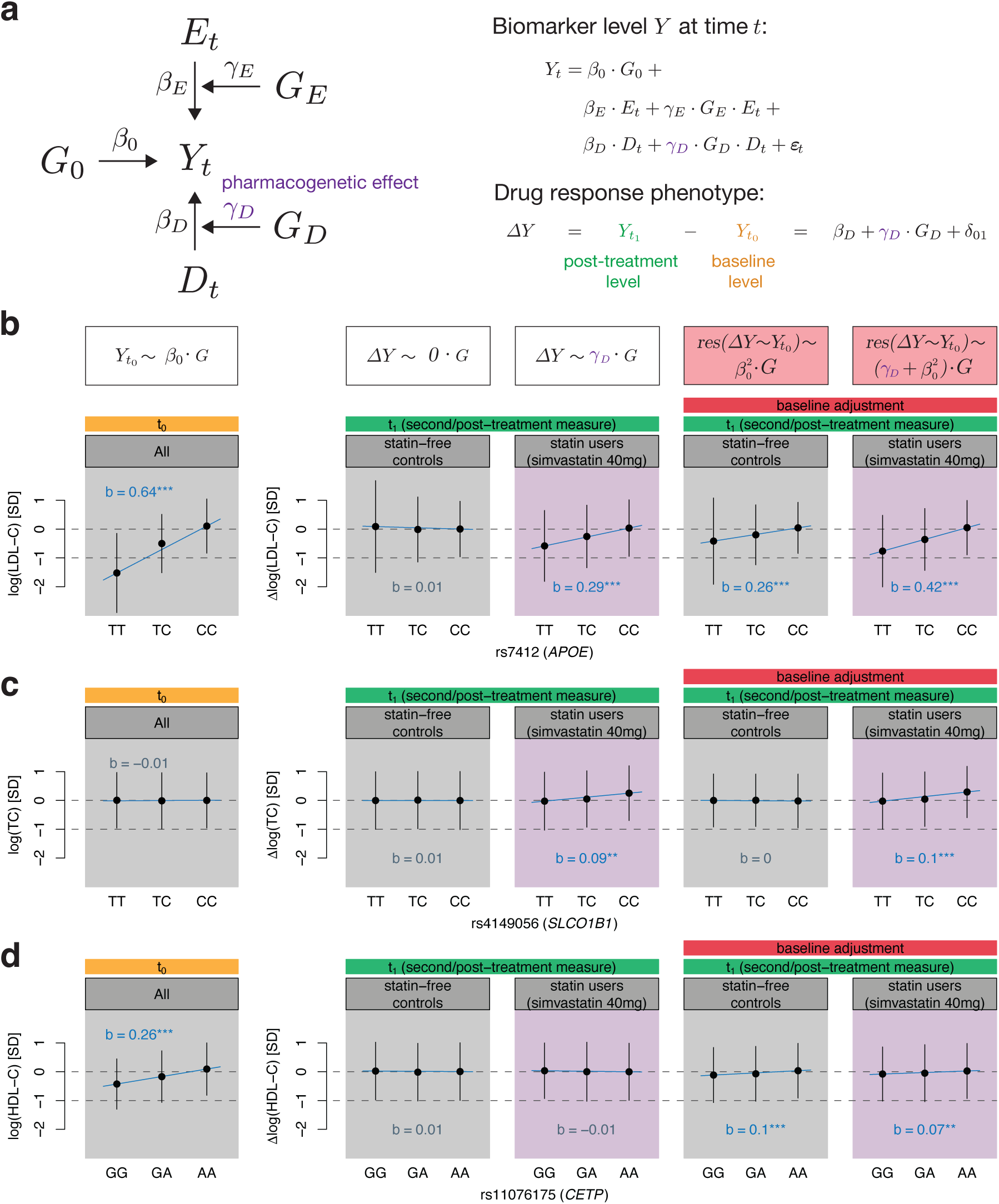
Modelling longitudinal changes of biomarker levels with (or without) treatment effect. **a** Biomarker levels *Y* at time *t* can be influenced by genetics *G*_0_, environment *E*, gene-environment interactions (*G_E_ · E*), drug status *D* and pharmacogenetic interactions (*G_D_ · D*). Drug response phenotypes modelled as the difference of post-treatment (t_1_) and baseline (t_0_) levels allow the estimation of the pharmacogenetic effect *γ_D_* through genetic regression analyses (Note S1). **b**-**d** Stratification at genetic variants that harbour pharmacogenetic *γ_D_* (**b**, **c**) and/or baseline *β*_0_ (**b**, **d**) genetic effects. Adjusting drug response or longitudinal change phenotypes for baseline induces a bias that scales with *β*_0_ (Note S1). Thus, variants with significant baseline effects spuriously associate with drug response phenotypes even if *γ_D_* is zero (**d**). Such measure of change, however, shows association in drug-naive individuals too. The baseline panel (t_0_) groups statin-free controls and statin users (simvastatin 40mg corresponding to the largest starting statin type-dose group), and shows their sex and age-adjusted standardized baseline level stratified by genotype. The following four panels (t_1_) show standardized longitudinal change (drug-naive individuals) and drug response phenotypes (statin users) adjusted for sex and age, once unadjusted (correct model) and adjusted (biased model) for baseline levels. Genotype regression co-efficients (denoted with *b*) with baseline lipid levels, longitudinal change and drug response phenotypes were derived through regression of the standardized outcome measures on the genotype dosage adjusted for sex and age as well as baseline levels if indicated. The significance level of the slope (*b*) is indicated by colour and stars where grey indicates a p-value *>* 0.05, blue a p-value *≤* 0.05, 2 stars a p-value *<* 1e-3 and 3 stars a p-value *<* 5e-8. Dots correspond to the mean and error bars to the standard deviation of covariate-adjusted baseline levels and drug response/longitudinal change phenotypes in each stratified group.

**Table 2:**
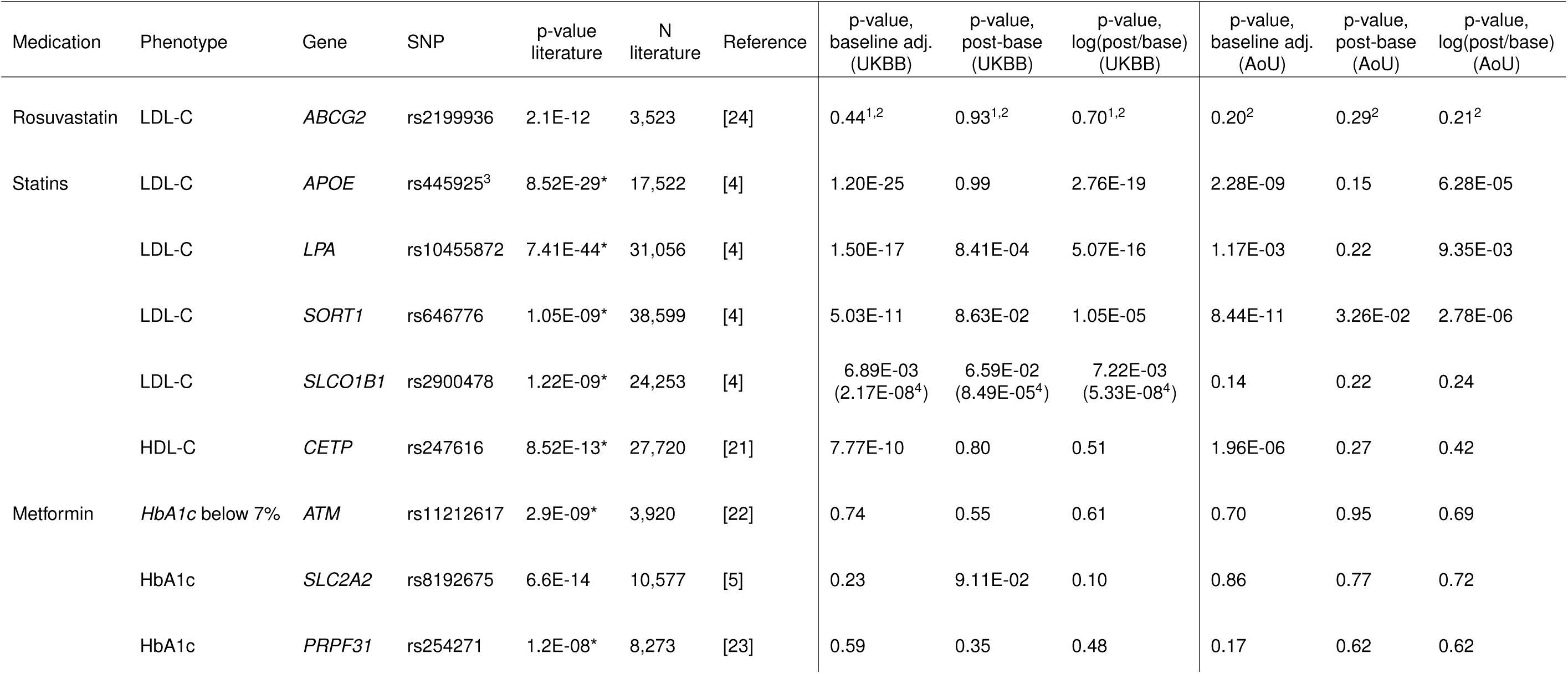
Genetic predictors of cardiometabolic drug response reported in the literature that were discovered or reproduced via genome-wide association study and passed a genome-wide significance threshold of 5e-8. Corresponding effect sizes and significance levels were retrieved from the EHR-derived genetic analyses in the discovery (UK Biobank) and replication (All of Us) cohort. baseline adj., absolute biomarker difference adjusted for baseline; post-base, absolute biomarker difference; log(post/base), logarithmic (relative) biomarker difference; UKBB, UK Biobank; AoU, All of Us. *results for baseline adjusted analysis ^1^results for LD-proxy rs45499402 (*r*^2^ = 1) ^2^all statins combined ^3^*r*^2^ with rs7412 = 0.64 ^4^results for LD-proxy rs4149056 (*r*^2^ = 0.87) and total cholesterol

GWAS of HbA1c-response to metformin identified *ATM* [22], *SLC2A2* [5] and *PRPF31* [23], none of which replicated in the EHR PGx GWAS (p-values *>* 0.05). While this could be a power issue given the lower sample sizes (4,119 and 3,641 in the UKBB and AoU, respectively), it should also be noted that none of the studies have reported the same locus twice and the *ATM* and *SLC2A2* loci were insignificant in the ACCORD clinical trial GWAS that was conducted later (p-value *>* 0.1) [23]. Although several loci have been found to influence blood pressure response to anti-hypertensives at a suggestive p-value threshold, no genome-wide significant hits have been reported [25].

### Rare variants have a modest impact

While common genetic variants have been assessed as predictors of drug response phenotypes in multiple studies, the impact of rare variation is less well known. Making use of sequencing data (WES and WGS in the UKBB and AoU, respectively), we conducted rare variant burden tests for all ten drug response phenotypes (Figure 1). We included missense and putative loss-of-function (LoF) variants with MAF *<* 1% in optimal kernel association tests (SKATO) [26]. After correcting for multiple testing (p-value *<* 0.05/18,983 = 2.63e-06), no gene passed the genome-wide significance threshold. When restricting the analysis to known pharmacogenes (66 very important (VIP) autosomal pharmacogenes defined by PharmGKB [27]), we found a significant association of rare variants in the *HLA-B* gene to influence relative LDL-C reduction following statin treatment (p-value = 4.62-04). This association could not be replicated in the AoU biobank (p-value = 0.24) indicating either a false positive association given the complexity of the *HLA* region and its sequencing difficulties, or a statistical power issue due to the lower sample size and the inclusion of different rare variants in the AoU.

### Modelling drug response and longitudinal change phenotypes

In the following, we will propose a longitudinal phenotype model to analyze drug response and longitudinal change phenotypes in an unbiased manner. Biomarker levels *Y* at time *t* can be modelled as follows (Figure 3a):

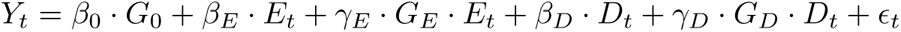

where *β*_0_ is the baseline genetic effect, *G* the genetics, *β_E_* the environmental effect, *E* the environment, *γ_E_* the gene-environment interaction effect, *D* the indicator of drug use, *β_D_* the drug effect and *γ_D_* the pharmacogenetic effect.

When modelling the drug response as the difference of post-treatment levels *Y_t_*_1_ and baseline levels *Y_t_*_0_, where the drug status is 1 and 0 at *t*_1_ and *t*_0_, respectively, the drug response phenotype simplifies to

(Note S1):

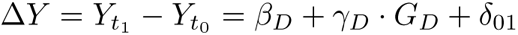

Thus, the pharmacogenetic effect can be estimated from genetic regression analyses on the biomarker difference. This expression also holds when modelling the logarithm of the biomarker level. However, adjusting biomarker differences for baseline levels induces a bias when genetic variants are also associated with baseline levels (*β*_0_ *̸*= 0). It can be shown that this bias is proportional to *β*^2^ (Note S1).

In Figure 3b-d, we depict genetic variants that either have a significant pharmacogenetic effect *γ_D_*, baseline effect *β*_0_ or both, and showcase how baseline adjustment can introduce a bias in genetic effect estimation. To this end, we compare genetic effect sizes of biomarker differences in medication-naive controls to those in statin users (simvastatin 40mg users who represent the largest starting statin type-dose group). The SNP rs7412 in the *APOE* region which is strongly associated to baseline levels (*β*_0_ = 0.636, p-value *<* 1e-300) also exhibits a pharmacogenetic effect (*γ_D_* = 0.294, p-value = 1.03e-18) while not being associated to longitudinal change in statin-free controls (p-value = 0.59; Figure 3b; Table S8-9). However, upon baseline adjustment, a significant genetic effect with longitudinal change is observed in drug naive individuals (b = 0.256, p-value = 4.83e-102) as well as a stronger association in statin users due to the *β*^2^ bias (b = 0.421, p-value = 4.12e-38). On the other hand, as rs4149056 (*SLCO1B1*) was not associated with total cholesterol baseline levels (p-value = 0.094), genetic effects remained similar between baseline adjusted and unadjusted results, evidencing the sole implication of *SLCO1B1* in pharmacokinetics (no significant association with longitudinal change and *γ_D_* = 0.094, p-value = 1.46e-07; Figure 3c). In contrast, the SNP rs11076175 in the *CETP* locus is strongly associated with HDL-C baseline levels (*β*_0_ = 0.263, p-value = 1.33e-307), but had no significant pharmacogenetic effect in the unbiased model (*γ_D_* = -0.007, p-value = 0.67). Upon baseline adjustment, strong associations with both longitudinal change and drug response were observed (p-values of 4.85e-33 and 3.2e-05, respectively; Figure 3d).

More generally, no genome-wide significant associations were found in (the correct) longitudinal biomarker progression GWAS in medication-naive individuals (Figures S9-10). However, upon base-line adjustment, striking similarities could be observed between drug response and longitudinal change GWAS (Figures S11-13; Tables S10-11). Additional genome-wide significant loci found in the baseline-adjusted GWAS included the *SORT1*/*CELSR2*/*PSRC1* locus in the LDL-C response as well as *CETP* in the HDL-C response to statin GWAS which also reached genome-wide significance in the longitudinal change GWAS in drug-naive participants.

### Polygenic risk scores as predictors of drug response

We assessed whether high PRS of the underlying biomarker contribute to better or worse drug response outcomes. High LDL-C PRS resulted in an increased absolute, albeit lower relative LDL-C reduction following statin treatment (b_abs_ = -0.085 mmol/L/SD PRS, p-value = 2.70e-36 and b_rel_ = 2.7%, p-value = 4.28e-36; Figure 4a; Table S12). These opposing effects of high PRS on absolute and relative drug efficacy were also reflected in the genetic correlations of drug response traits with baseline traits. While the absolute LDL-C genetic difference was negatively correlated to LDL-C baseline levels (*r_g_* = -1.08, 95%CI = [-1.52, -0.65]), the genetic correlation with the relative LDL-C difference was positive, although not significant (*r_g_* = 0.195, 95%CI = [-0.02, 0.42]; Figure S14; Table S13). Association results between TC PRS and TC response to statins were highly significant (p-value *<* 5.39e-19) and directionally concordant with LDL-C results. Nominally significant results between biomarker PRS and drug response phenotypes were found for high HDL-C PRS and increased HDL-C levels following statin treatment (b_abs_ = 0.003 mmol/L/SD PRS, p-value = 0.019), high HbA1c PRS decreasing relative change following metformin treatment (b_rel_ = 1.02%/SD PRS, p-value = 6.79e-03) and high SBP PRS increasing SBP reduction following ACEi treatment (b_abs_ = -0.53 mmHg/SD PRS, p-value = 0.035; Table S12). As for genetic association analyses, care has to be taken not to adjust for baseline levels as this can reverse directionality due to PRS affecting both baseline and biomarker reduction: adjusting the LDL-C reduction for LDL-C baseline levels suggests that high PRS decreases treatment efficacy (b_abs_ = 0.14 mmol/L/SD PRS, p-value = 2.74e-141). As a control experiment, a high PRS led to a nominally significant increase in LDL-C in statin-free controls at the second measure (b_abs_ = 0.006 mmol/L/SD PRS, p-value = 0.045), which upon baseline adjustment inflated to the same value as in statin users (b_abs_ = 0.14 mmol/L/SD PRS, p-value *<* 1e-300).

**Figure 4:**
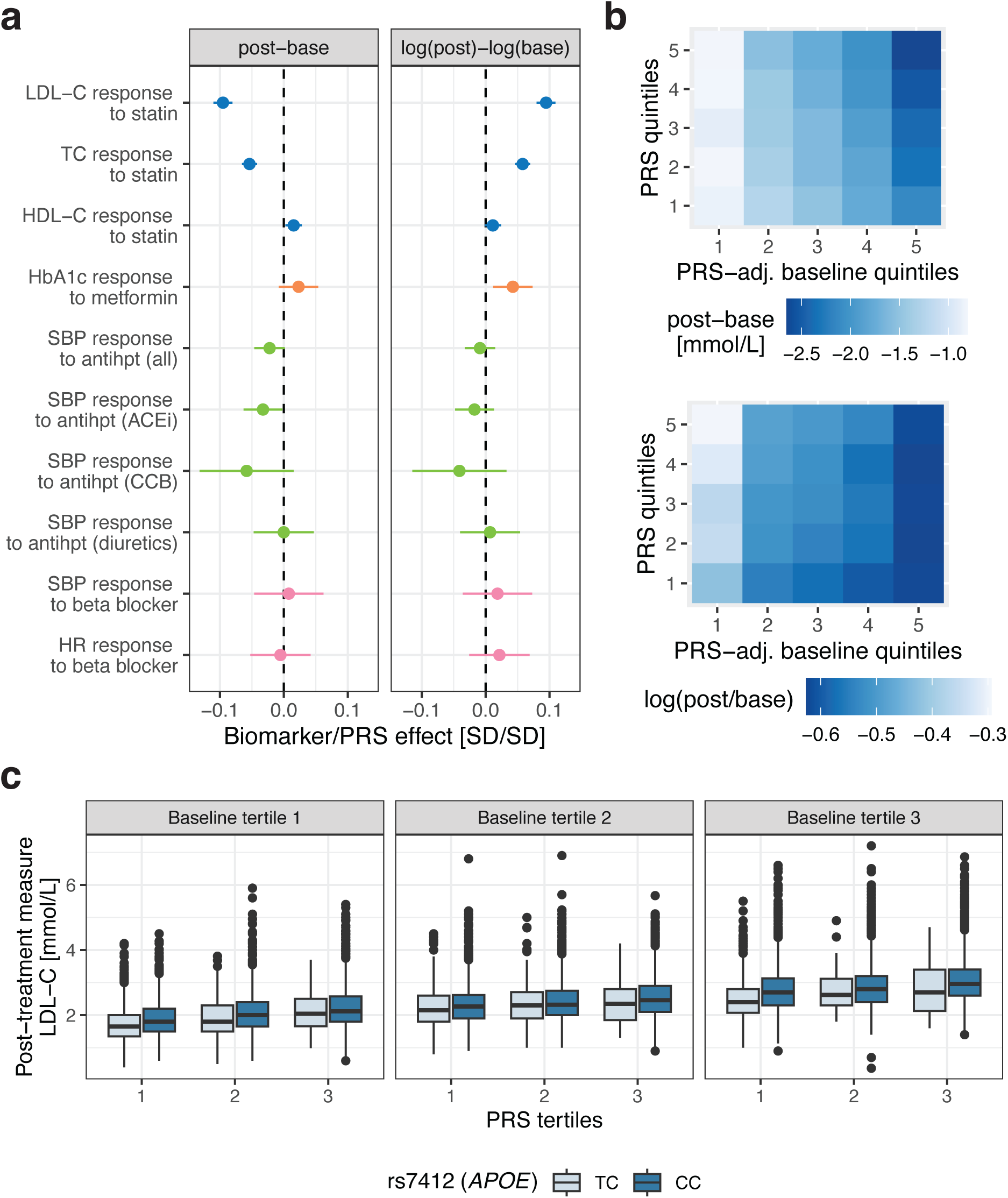
Drug response phenotype associations with PRS. **a** Drug response associations with PRS calculated for the absolute (post-base) and logarithmic relative (log(post)-log(base)) biomarker difference colour-coded by the drug. Standardized effect sizes (biomarker/PRS effect) correspond to an SD change for 1SD increase in PRS. A negative sign means that increased PRS increases treatment efficacy (i.e., larger biomarker difference compared to low PRS). All associations are adjusted for sex, age and drug-specific covariates. **b** Statin users stratified by 1) LDL-C baseline levels adjusted for LDL-C PRS and 2) LDL-C PRS quintiles with each tile showing the average LDL-C biomarker response (top: absolute, bottom: relative difference). Darker blue values correspond to stronger biomarker reductions. **c** Statin users stratified by 1) LDL-C baseline levels, 2) LDL-C PRS and 3) rs7412 genotype (individuals with the TT genotype are omitted as their sample size was too low). Boxes bound the 25th, 50th (median, centre), and the 75th quantile of LDL-C post-treatment measures. Whiskers range from minima (Q1 – 1.5*IQR) to maxima (Q3 + 1.5*IQR) with points above or below representing potential outliers.

Although PRS can serve as a predictor of drug response, starting baseline levels remain the best predictor of post-treatment levels (Figure 4b-c). To disentangle the effect of genetics and environment on drug response, we adjusted baseline levels for PRS as the resulting PRS-adjusted baseline levels should more closely reflect the environmental component. PRS-adjusted baseline levels explained 42.2% and 11.3% of the variance of absolute and relative LDL-C difference, respectively. Increased LDL-C PRS increased LDL-C reduction which was most pronounced at high PRS-adjusted baseline levels (Figure 4b). Conversely, high LDL-C PRS decreased relative LDL-C reduction which was more apparent at low PRS-adjusted baseline levels. By integrating the PRS, the explained variance increased from 42.2 to 43.3% and from 11.3 to 12.1% for absolute and relative LDL-C differences, respectively.

In Figure 4c, we highlight how additional stratification by drug response genetic signals can improve prediction accuracy for post-treatment LDL-C levels following statin initiation (Table S14). Additional stratification by the *APOE* genotype, the top signal in the LDL-C response GWAS, increased the explained variance of the relative reduction to 12.4% compared to 12.1% for adjusted baseline and LDL-C PRS predictors alone. Post-treatment levels remain higher for individuals with high PRS despite greater absolute LDL-C reductions.

## Discussion

In this study, we demonstrate the value of biobanks coupled with EHRs to study the genetics of cardiometabolic disease medications. We conducted discovery in the UKBB and replication analyses in the AoU, and assessed the impact of common and rare variations on drug efficacy. We show that signals from EHR-derived PGx GWAS are concordant with those observed in the literature and present a theoretical framework to model drug response and longitudinal phenotypes.

Overall, we found only a few genetic variants to influence cardiometabolic drug response in line with other studies that often identified only a few or even no genome-wide significant signals [6, 8, 28]. A review on drug efficacy GWAS reported that only 15% of drugs exhibit robust gene-treatment interactions [2], the extent of which largely depends on the drug’s mode of action. While we could identify *LDLR* and *ZNF800* as novel loci, sample sizes remain too low to have a definite answer on whether low numbers of genetic predictors are a consequence of limited statistical power or a lack of genetic influence on drug response which would be corroborated by low and often insignificants heritability estimates (Table S13). While inter-individual variability in drug response could also depend on rare variants, we could not identify robust rare variant associations in this study.

As similar study designs have used differing GWAS models to estimate pharmacogenetic effects, we elucidated the theory of modelling biomarker differences and showed how baseline adjustment can induce biases for genetic variants associated with baseline levels. For unbiased pharmacogenetic effect estimation, both absolute and logarithmic relative biomarker differences can be assessed. Whether absolute or relative reductions are most determining in lowering the risk of associated clinical diseases may be dependent on the studied medication-biomarker pair. If biomarker levels are linearly associated with the disease risk, absolute change is more relevant; however, if this relationship is exponential or quadratic, then relative change is of more importance. As to pharmacogenetic interactions, it is a priori unknown on which scale genetic variants act linearly, evidencing the usefulness of testing both absolute and relative biomarker changes.

In line with other studies, we found that high PRS of the underlying biomarker can lead to increased absolute, although lower relative biomarker reductions. A recent study showed that sulfonylureas therapy was more effective in participants with higher T2D PRS with findings replicated in a separate cohort [29]. Using RCT data, high coronary heart disease (CHD) genetic risk was found to be associated with increased CHD risk, although the comparison between controls and treated participants revealed that relative risk reductions were higher among individuals with a high PRS, suggesting that this group benefited the most from lipid-lowering therapy [30, 31, 32, 33]. Given the complexity of genetics affecting both baseline biomarker levels and disease risk as well as reductions thereof, disentangling whether genetic or environmental factors can be easier alleviated by medication requires careful considerations as adjusting for baseline levels induces genetic biases. While we found strong evidence for a higher genetic burden to increase low-density and total cholesterol reductions, this effect artificially reverses when adjusting for baseline levels. While RCT data with a control arm remains the gold standard for studying such complex interactions, large biobank data also allow the construction of (non-randomized) control groups. Both for variant-level and PRS association analyses, we demonstrate how drug response and disease progression genetics seemingly overlap when adjusting for baseline and how these base-line genetics signals disappear in the control group upon applying the correct longitudinal change model.

Our study has several limitations. First, we rely on data from EHRs to derive before and after treatment biomarker levels, and thus cannot exclude the possibility that individuals were already on medication before the first recorded prescription. Second, despite a large fraction of individuals with medication records in the biobanks, final PGx cohort sample sizes are limited by the number of participants on a certain medication and further reduced due to incomplete or missing data. Of the ∼65,000 participants with a statin prescription in the UKBB, 63% could not be considered for the LDL-C response analysis because of missing baseline and/or post-treatment measures. Third, polypharmacy has only been taken into account within and not across medication groups. Even within, especially for antihypertensives where frequent changes in medication regimen occur, it can be difficult to determine appropriate filtering and covariate strategies to study individual drug classes as sample sizes are too low when restricting the analysis to individuals taking antihypertensives from a single class (i.e., stringent filtering strategy). Fourth, our analysis focused on continuous biomarkers and not on clinical events. LDL-C, SBP, HR and HBA1c merely serve as surrogate endpoints of CHD and T2D events, and the genetic interplay with drug efficacy may be different when assessing hard clinical endpoints. Finally, we rely on observational data to draw conclusions about drug efficacy. Although, we contrast the results with control analyses on longitudinal biomarker change, control and medication groups were not defined randomly and by definition have markedly different disease profiles.

To conclude, we show that EHRs enable new opportunities to study drug response and reveal the complex contribution of genetic and environmental components to drug efficacy. While we find that the influence of common and rare genetic variants on drug response is relatively low, larger sample sizes will be needed to capture the full extent.

## Methods

### Study population

The UK Biobank is a prospective study of ∼500,000 participants of whom 45% (N *≈* 230,000) are linked to the primary care data of the United Kingdom’s National Health System [16]. The primary care resource contains longitudinal data of GP prescription records (datafield #42039) and GP clinical event records (datafield #42040) encoded through the British National Formulary (BNF), National Health Service (NHS) dictionary of medicines and devices (DM+D), Read V2 and Clinical Terms Version 3 (CTV3) codes and are available up to 2016 or 2017 (depending on the data supplier). Analyses were conducted on individuals of white British ancestry, with no excessive number of relatives and differing reported and inferred gender, excluding participants who have withdrawn their consent (UKBB Sample-QC #531; N *≈* 200,000).

### Study design and drug response phenotypes

We derived drug response phenotypes for the following cardiometabolic medication-phenotype pairs: statin-lipids (LDL-C, HDL-C, TC), metformin-HbA1c, antihypertensive-SBP (by antihypertensive class and all classes combined), beta blocker-SBP and beta blocker-HR. For each drug response phenotype, we considered stringent and lenient filtering scenarios which differed by regularity in prescription pattern, pre-treatment and post-treatment time windows as well as handling of treatment changes (e.g. dose change) and concomitant medication (e.g. add-on therapy). In Figure S1 and Table S3, we outline the different QC filters applied to each scenario. To further increase the number of available clinical measures, we added measures from the initial and repeated assessment visits with their respective time stamps to the pool of longitudinal data (LDL-C: #30780, HDL-C: #30760, TC: #30690, HbA1c: #30750, SBP: #4080, HR: #102). Read V2 and CTV3 codes encoding these variables in the primary care data are listed in Table S1 (see Note S2 for HbA1c unit conversion). Baseline measures were taken three months (stringent filtering) or up to a year (lenient filtering) before treatment initiation and 7 days after, either as the closest measure to treatment start or an average of all available measures during the pre-treatment period. The post-treatment period was defined as 6 months after medication start up to 1.5 (stringent) and 2 (lenient) years after, and either the closest measure to treatment start or an average of all available measures during the post-treatment period were taken. Consequently, we derived drug response phenotypes for four scenarios: stringent filtering-single measure, stringent filtering-average measures, lenient filtering-single measure, and lenient filtering-average measures.

To determine medication regimens (medication start, treatment changes, prescription regularity), we first extracted all available prescriptions for each broader medication class (lipid-regulating, antidiabetic including insulin, and antihypertensives; BNF and Read V2 codes in Table S2). We then selected individuals with entries of the medication of interest (primary medication) and omitted individuals taking medications other than the primary medication of the same class within a year of initiating the primary medication. Note that in the analysis of beta blockers for SBP we excluded individuals taking other antihypertensives, whereas such individuals were not excluded for HR since their effect on HR is much weaker than for beta blockers [34]. In the lenient filtering scenarios, we considered exceptions to this rule such as metformin being an add-on therapy to sulfonylureas, with sulfonylureas treatment being a covariate. Allowed scenarios for add-on therapy for antihypertensive are shown in Figure S2. Individuals taking primary medications in combination with a medication of the same class (e.g. statins in combination with ezetimibe) were filtered out (Note S3). If multiple drugs corresponded to a medication class (e.g. different statin types), we included all drugs taken by at least 20 individuals. When BNF codes were truncated to miss the drug ingredient, we extracted them by matching drug names and brand names in the drug description. Likewise, dosage information was retrieved from the description using regular expressions [13].

In Table S4, we show the study characteristics of the individuals in each drug response phenotype cohort. Furthermore, bar plots in Figure S3 show the number of individuals after each QC step. The different QC steps were as follows: i) available baseline and post-treatment measures, ii) presence of a primary care record other than baseline/primary medication at least two years before the medication start to avoid falsely considering a change to a new health care provider as a first prescription, iii) presence of a prescription part of the broader medication class after post-treatment measure, iv) drug change between medication start and post-treatment measure, v) regular prescriptions proxying drug adherence and vi) minimum baseline level (e.g. LDL-C *≥* 2 mmol/L). We only considered cohorts with more than 500 individuals for GWAS analyses.

### GWAS

In the genetic association analyses, we define the drug response phenotype as either the absolute (*Y_t_*_1_ *− Y_t_*_0_) or the logarithmic relative (*log*(*Y_t_*_1_) *− log*(*Y_t_*_0_) = *log*(*Y_t_*_1_ */Y_t_*_0_)) difference of post-treatment *Y_t_*_1_ and baseline *Y_t_*_0_ levels. This difference was then adjusted for study-specific covariates including sex, age at the time of medication start, time between medication start and post-treatment measure, drug type and dose if applicable and the first 20 principal components (Table S3). Importantly, the difference was not adjusted for baseline levels as this can induce a bias for genetic variants that are associated with baseline (Note S1).

GWAS analyses were conducted using REGENIE (v3.2.6) which accounts for sample relatedness [35]. REGENIE first fits a whole-genome regression model (step 1) before testing each SNP in a leave-one-chromosome-out (LOCO) scheme (step 2). In step 1, genotyped SNPs were filtered as follows using PLINK2 [36]: minor allele frequency (MAF) *≥* 0.01, Hardy-Weinberg equilibrium p-value *≥* 1e-15, genotyping rate *≥* 0.99, not present in high linkage disequilibrium (LD) regions [37], not involved in inter-chromosomal LD [35] and passing LD pruning at *r*^2^ *<* 0.9 with a window size of 1,000 markers and a step size of 100 markers which resulted in 424,544 SNPs included in step 1. In step 2, variants imputed by the Haplotype Reference Consortium panel with a MAF *≥* 0.05 were tested (up to 5.5 million markers depending on phenotype sample size). Individuals with missing genetic data and/or not passing genetic QC were excluded from the analysis. Independent signals were defined as *r*^2^ *<* 0.001 and clumping was performed using PLINK and the UK10K reference panel [38].

### Rare variant analysis

Rare variant analyses were conducted using REGENIE (v3.2.9). Phenotype definitions and covariates (Table S3) were the same as in the GWAS analyses, except that biomarker differences were transformed by inverse quantile normalization to decrease the chance of false positives. Following step 1 whole genome regression (Method section: GWAS), we performed rare variant burden tests using optimal kernel association tests (SKATO) in step 2 [26]. Masks were constructed from rare variants (MAF *<* 0.01) including missense and putative LoF variants, and REGENIE SKATO tests were computed with default parameters. Variant annotations and gene set definitions were derived following the original quality functionally equivalent (OQFE) protocol and provided on the UK Biobank DNAnexus research analysis platform [39]. Burden tests were then conducted on OQFE WES data (#23158) [39]. Genes classified as very important pharmacogenes (VIPs) were downloaded from the PharmGKB gene annotations (April 5, 2023 version)[27].

### Replication in the All of Us biobank

The All of Us research program is a prospective cohort recruiting up to 1 million participants [17]. Replication analyses were conducted in the current release (v7) in which genotype data were available for ∼310,000 and WGS data for ∼250,000 individuals. In the AoU database, the Observational Medical Outcomes Partnership (OMOP) Common Data Model (CDM) is used for standardized vocabularies and harmonized data representations. Medication records were retrieved based on concept ID codes from the RxNorm vocabulary and phenotypes from the SNOMED vocabulary. Replication analyses were restricted to lipid response to statins and HbA1c response to metformin for which genome-wide significant signals were obtained either in the UKBB analyses or reported in the literature.

Similarly to the UKBB, we extracted medication records by starting from the broader medication class (lipid modifying agents (concept id 21601853) and drugs used in diabetes (concept id 21600712)) which were then classified into primary medications (statins (concept id 21601855) and metformin (concept id 1503297)), combination therapies (lipid modifying agents, combinations (concept id 21601898), blood glucose lowering drugs, combination (concept id 21600765) and sulfonylureas (concept id 21600749)) and related medication from the same class. Dose information was extracted from the drug concept entries using regular expression or imputed by the median dose of the drug in question when not available. Phenotypes were extracted based on the following ancestor concept IDs: LDL-C (3028437), HDL-C (3007070), TC (3027114) and HbA1c (3004410). Only measures with available units and values in the plausible range were retained (Table S1). While lipid measures were recorded as mmol/L and primarily as mmol/mol for HbA1c in the UKBB (Note S2), units were mg/dL and % for lipids and HbA1c, respectively, which we left unconverted. Following the extraction of longitudinal medication and biomarker measures, we followed the same QC steps as in the UKBB by applying the lenient filtering strategy with average baseline and post-treatment measures (Figure S1). Drug prescription regularity was found to be lower in the AoU, likely because drug prescriptions are only recorded from participating EHR sites. As a consequence, we lowered the drug regularity QC parameter and required a single prescription between medication start and post-treatment measures (QC9, Figure S1). Cohort characteristics and reason for removal are reported in Table S7 and Figure S8, respectively.

#### GWAS

GWAS analyses were conducted using REGENIE (v3.2.4). For step 1, we used genotyped SNPs and filtered them as follows using PLINK2: autosomal SNPs, MAF *≥* 0.01, Hardy-Weinberg equilibrium p-value *≥* 1e-15, genotyping rate *≥* 0.99, not present in high linkage disequilibrium (LD) regions [37] and passing LD pruning at *r*^2^ *<* 0.9 with a window size of 1,000 markers and a step size of 100 markers which resulted in 238,888 SNPs. The first 20 PCs were computed on the same set of SNPs using the FastPCA algorithm implemented in PLINK2 [40]. In step 2, we used WGS data from the Allele Count/Allele Frequency (ACAF) threshold callset to test associations between the genotypes of interest and drug response phenotypes.

#### Rare variant analysis

We conducted SKATO analyses on rare variants from the exon regions using REGENIE (v3.2.4) with step 1 being the same as in the GWAS. Variant annotations and gene set definitions were extracted from the Variant Annotation Table (VAT) provided by the AoU. Missense variants and putative LoF variants defined as stop gained, frameshift, splice donor and splice acceptor with MAF *<* 0.01 were included in the burden tests.

### PRS and genetic correlations

We calculated PRS for UKBB participants with the PGS Catalog Calculator [41, 42] using pre-calculated genetic effect sizes from the PGS Catalog [43]: LDL-C, PGS002150; HDL-C, PGS002172; TC, PGS002108; HbA1c, PGS002171; SBP, PGS002228; HR, PGS002193. We then calculated the associations between PRS and the absolute and (logarithmic) relative biomarker difference adjusted for sex, age and drug-specific covariates (Table S3).

We calculated genetic correlations between traits using the GenomicSEM R package (v0.0.5c) [44]. Trait GWAS summary statistics were obtained from the following consortia: LDL-C, HDL-C and TC from the Global Lipids Genetics Consortium [45] (N up to 1,320,016; European ancestry), HbA1c from the UKBB (#30750, N = 344,182), SBP from a meta-analysis of the UKBB and the International Consortium of Blood Pressure [46] (N up to 757,601) and HR from the UKBB (#102, N = 340,162) where the UKBB GWAS summary statistics came from Neale’s lab (http://www.nealelab.is/uk-biobank).

### Longitudinal biomarker change GWAS

We conducted biomarker change GWAS in individuals part of the primary care data that did not have any drug prescription indicated for the investigated disease/surrogate endpoint (i.e., broad medication class, Table S2). All participants in this set with two available measures spaced between 6 months and 3 years which corresponds to the minimum and maximum allowed time interval between baseline and post-treatment measures were included. GWAS analyses were conducted analogous to the drug response GWAS, replacing baseline with first and post-treatment with second phenotype measure. We used the same covariates as in the corresponding drug response cohorts omitting drug-specific variables (Table S3).

## Data Availability

All GWAS summary statistics produced in the present work will be available in the GWAS Catalog upon publication.

## Data availability

Genetic and phenotypic data from the UK Biobank and All of Us Resource are available to approved researchers.

British National Formulary (BNF), National Health Service (NHS) dictionary of medicines and devices (DM+D), Read V2 and Clinical Terms Version 3 (CTV3) vocabularies encoding the UK Biobank primary care records, https://biobank.ndph.ox.ac.uk/showcase/refer.cgi?id=592.

ATHENA – OHDSI vocabulary repository for RxNorm (drug) and SNOMED (phenotype) concept IDs, https://athena.ohdsi.org/.

Polygenic risk score genetic effect sizes (UK Biobank), https://www.pgscatalog.org/publication/PGP000263/.

Lipid GWAS summary statistics from the Global Lipids Genetics Consortium, https://csg.sph.umich. edu/willer/public/glgc-lipids2021/.

Systolic blood pressure GWAS summary statistics from the UK Biobank and the International Consortium of Blood Pressure meta-analysis, https://www.ebi.ac.uk/gwas/publications/30224653.

Neale’s lab GWAS summary statistics (UK Biobank), http://www.nealelab.is/uk-biobank.

UK10K individual-level data are available upon request, https://www.uk10k.org/data_access.html.

All GWAS and rare variant burden test summary statistics will be available in the GWAS Catalog upon publication.

## Code availability

GWAS calculations were performed with REGENIE (v3.2.6) which is available at https://github.com/ rgcgithub/regenie. PLINK2 is available at https://www.cog-genomics.org/plink/2.0/. PRS were calculated with the PGS Catalog Calculator (v2.0) available at https://github.com/PGScatalog/pgsc_calc. Genetic correlations were calculated with the GenomicSEM R package (v0.0.5c) available at https://github.com/GenomicSEM/GenomicSEM. All codes used in this analysis are available on GitHub (https://github.com/masadler/PGxEHR).

## Authors’ contributions

MCS and ZK conceived and designed the study. MCS performed statistical analyses in the UK Biobank. AA conducted replication analyses in the All of Us research program under the supervision of MCS and RBA. CCB computed PRS on UK Biobank participants. DMR provided guidance on analyzing rare variants from sequencing data. ZK supervised all statistical analyses. All the authors contributed by providing advice on the interpretation of results. MCS and ZK drafted the manuscript. All authors read, approved, and provided feedback on the final manuscript.

## Acknowledgements

This work was supported by the Swiss National Science Foundation (310030 189147) to ZK. RBA is supported by NIH HG010615, Chan Zuckerberg Biohub, and Burroughs Wellcome Fund. MCS would like to thank the Fulbright Program for funding her research stay at Stanford University where this research work has started under the co-supervision of RBA. This research has been conducted using the UK Biobank Resource under Application Number 16389. LD was calculated based on the UK10K data resource (EGAD00001000740, EGAD00001000741). Computations were performed on the Urblauna cluster of the University of Lausanne. We also would like to acknowledge the participants and investigators of the UK Biobank and All of Us study. We thank Greg McInnes for his help and support in analyzing medication prescription data in the UK Biobank and Ewan Pearson for his advice in defining drug response phenotypes from the UK Biobank primary care data.

## References

[1] Gregory McInnes, Sook Wah Yee, Yash Pershad, and Russ B Altman. Genomewide association studies in pharmacogenomics. Clinical Pharmacology & Therapeutics, 110(3):637–648, 2021.

[2] Matthew R Nelson, Toby Johnson, Liling Warren, Arlene R Hughes, Stephanie L Chissoe, Chun-Fang Xu, and Dawn M Waterworth. The genetics of drug efficacy: opportunities and challenges. Nature Reviews Genetics, 17(4):197–206, 2016.

[3] Munir Pirmohamed. Pharmacogenomics: Current status and future perspectives. Nature Reviews Genetics, pages 1–13, 2023.

[4] Iris Postmus, Stella Trompet, Harshal A Deshmukh, Michael R Barnes, Xiaohui Li, Helen R Warren, Daniel I Chasman, Kaixin Zhou, Benoit J Arsenault, Louise A Donnelly, et al. Pharmacogenetic meta-analysis of genome-wide association studies of ldl cholesterol response to statins. Nature communications, 5(1):5068, 2014.

[5] Kaixin Zhou, Sook Wah Yee, Eric L Seiser, Nienke Van Leeuwen, Roger Tavendale, Amanda J Bennett, Christopher J Groves, Ruth L Coleman, Amber A Van Der Heijden, Joline W Beulens, et al. Variation in the glucose transporter gene slc2a2 is associated with glycemic response to metformin. Nature genetics, 48(9):1055–1059, 2016.

[6] Adem Y Dawed, Andrea Mari, Andrew Brown, Timothy J McDonald, Lin Li, Shuaicheng Wang, Mun-Gwan Hong, Sapna Sharma, Neil R Robertson, Anubha Mahajan, et al. Pharmacogenomics of glp-1 receptor agonists: a genome-wide analysis of observational data and large randomised controlled trials. The Lancet Diabetes & Endocrinology, 11(1):33–41, 2023.

[7] Erika Salvi, Zhiying Wang, Federica Rizzi, Yan Gong, Caitrin W McDonough, Sandosh Padmanabhan, Timo P Hiltunen, Chiara Lanzani, Roberta Zaninello, Martina Chittani, et al. Genome-wide and gene-based meta-analyses identify novel loci influencing blood pressure response to hydrochlorothiazide. Hypertension, 69(1):51–59, 2017.

[8] Sonal Singh, Helen R Warren, Timo P Hiltunen, Caitrin W McDonough, Nihal El Rouby, Erika Salvi, Zhiying Wang, Tatiana Garofalidou, Frej Fyhrquist, Kimmo K Kontula, et al. Genome-wide meta-analysis of blood pressure response to *β*1-blockers: results from icaps (international consortium of antihypertensive pharmacogenomics studies). Journal of the American Heart Association, 8(16):e013115, 2019.

[9] Caitrin W McDonough, Helen R Warren, John R Jack, Alison A Motsinger-Reif, Nicole D Armstrong, Joshua C Bis, John S House, Sonal Singh, Nihal M El Rouby, Yan Gong, et al. Adverse cardiovascular outcomes and antihypertensive treatment: A genome-wide interaction meta-analysis in the international consortium for antihypertensive pharmacogenomics studies. Clinical Pharmacology & Therapeutics, 110(3):723–732, 2021.

[10] William S Bush, David R Crosslin, A Owusu-Obeng, John Wallace, Berta Almoguera, Melissa A Basford, Suzette J Bielinski, David S Carrell, John J Connolly, Dana Crawford, et al. Genetic variation among 82 pharmacogenes: The PGRNseq data from the eMERGE network. Clinical Pharmacology & Therapeutics, 100(2):160–169, 2016.

[11] Chiara Auwerx, Marie C Sadler, Alexandre Reymond, and Zoltán Kutalik. From pharmacogenetics to pharmaco-omics: Milestones and future directions. Human Genetics and Genomics Advances, 2022.

[12] Tönis Tasa, Kristi Krebs, Mart Kals, Reedik Mägi, Volker M Lauschke, Toomas Haller, Tarmo Puu-rand, Maido Remm, Tönu Esko, Andres Metspalu, et al. Genetic variation in the Estonian population: pharmacogenomics study of adverse drug effects using electronic health records. European Journal of Human Genetics, 27(3):442–454, 2019.

[13] Gregory McInnes and Russ B Altman. Drug response pharmacogenetics for 200,000 uk biobank participants. In BIOCOMPUTING 2021: Proceedings of the Pacific Symposium, pages 184–195. World Scientific, 2020.

[14] Mustafa Adnan Malki, Adem Y Dawed, Caroline Hayward, Alex Doney, and Ewan R Pearson. Utilizing large electronic medical record data sets to identify novel drug–gene interactions for commonly used drugs. Clinical Pharmacology & Therapeutics, 110(3):816–825, 2021.

[15] Tuomo Kiiskinen, Pyry Helkkula, Kristi Krebs, Juha Karjalainen, Elmo Saarentaus, Nina Mars, Arto Lehisto, Wei Zhou, Mattia Cordioli, Sakari Jukarainen, et al. Genetic predictors of lifelong medication-use patterns in cardiometabolic diseases. Nature Medicine, 29(1):209–218, 2023.

[16] Clare Bycroft, Colin Freeman, Desislava Petkova, Gavin Band, Lloyd T Elliott, Kevin Sharp, Allan Motyer, Damjan Vukcevic, Olivier Delaneau, Jared O’Connell, et al. The UK Biobank resource with deep phenotyping and genomic data. Nature, 562(7726):203–209, 2018.

[17] The All of Us Research Program Investigators. The “All of Us” research program. The New England Journal of Medicine, 381(7):668–676, 2019.

[18] Jemma C Hopewell, Sarah Parish, Alison Offer, Emma Link, Robert Clarke, Mark Lathrop, Jane Armitage, Rory Collins, and MRC/BHF Heart Protection Study Collaborative Group. Impact of common genetic variation on response to simvastatin therapy among 18 705 participants in the heart protection study. European heart journal, 34(13):982–992, 2013.

[19] Jaideep Patel, H Robert Superko, Seth S Martin, Roger S Blumenthal, and Lisa Christopher-Stine. Genetic and immunologic susceptibility to statin-related myopathy. Atherosclerosis, 240(1):260– 271, 2015.

[20] Elliot Sollis, Abayomi Mosaku, Ala Abid, Annalisa Buniello, Maria Cerezo, Laurent Gil, Tudor Groza, Osman Günesş, Peggy Hall, James Hayhurst, et al. The NHGRI-EBI GWAS Catalog: knowledgebase and deposition resource. Nucleic Acids Research, 51(D1):D977–D985, 2023.

[21] Iris Postmus, Helen R Warren, Stella Trompet, Benoit J Arsenault, Christy L Avery, Joshua C Bis, Daniel I Chasman, Catherine E de Keyser, Harshal A Deshmukh, Daniel S Evans, et al. Meta-analysis of genome-wide association studies of HDL cholesterol response to statins. Journal of medical genetics, 53(12):835–845, 2016.

[22] Lorna W Harries, Andrew T Hattersley, Alex SF Doney, Helen Colhoun, Andrew D Morris, Calum Sutherland, D Grahame Hardie, Leena Peltonen, Mark I McCarthy, et al. Common variants near ATM are associated with glycemic response to metformin in type 2 diabetes. Nature genetics, 43(2):117–120, 2011.

[23] Daniel M Rotroff, Sook Wah Yee, Kaixin Zhou, Skylar W Marvel, Hetal S Shah, John R Jack, Tammy M Havener, Monique M Hedderson, Michiaki Kubo, Mark A Herman, et al. Genetic variants in CPA6 and PRPF31 are associated with variation in response to metformin in individuals with type 2 diabetes. Diabetes, 67(7):1428–1440, 2018.

[24] Daniel I Chasman, Franco Giulianini, Jean MacFadyen, Bryan J Barratt, Fredrik Nyberg, and Paul M Ridker. Genetic determinants of statin-induced low-density lipoprotein cholesterol reduction: the Justification for the Use of Statins in Prevention: an Intervention Trial Evaluating Rosuvastatin (JUPITER) trial. Circulation: Cardiovascular Genetics, 5(2):257–264, 2012.

[25] Gustavo H Oliveira-Paula, Sherliane C Pereira, Jose E Tanus-Santos, and Riccardo Lacchini. Pharmacogenomics and hypertension: current insights. Pharmacogenomics and personalized medicine, pages 341–359, 2020.

[26] Seunggeun Lee, Michael C Wu, and Xihong Lin. Optimal tests for rare variant effects in sequencing association studies. Biostatistics, 13(4):762–775, 2012.

[27] Michelle Whirl-Carrillo, Rachel Huddart, Li Gong, Katrin Sangkuhl, Caroline F Thorn, Ryan Whaley, and Teri E Klein. An evidence-based framework for evaluating pharmacogenomics knowledge for personalized medicine. Clinical Pharmacology & Therapeutics, 110(3):563–572, 2021.

[28] Cong Zhang, Konstantin Shestopaloff, Benjamin Hollis, Chun Hei Kwok, Claudia Hon, Nicole Hartmann, Chengeng Tian, Magdalena Wozniak, Luis Santos, Dominique West, et al. Response to anti-IL17 therapy in inflammatory disease is not strongly impacted by genetic background. The American Journal of Human Genetics, 2023.

[29] Josephine H Li, Lukasz Szczerbinski, Adem Y Dawed, Varinderpal Kaur, Jennifer N Todd, Ewan R Pearson, and Jose C Florez. A polygenic score for type 2 diabetes risk is associated with both the acute and sustained response to sulfonylureas. Diabetes, 70(1):293–300, 2021.

[30] Jessica L Mega, Nathan O Stitziel, J Gustav Smith, Daniel I Chasman, Mark J Caulfield, James J Devlin, Francesco Nordio, Craig L Hyde, Christopher P Cannon, Frank M Sacks, et al. Genetic risk, coronary heart disease events, and the clinical benefit of statin therapy: an analysis of primary and secondary prevention trials. The Lancet, 385(9984):2264–2271, 2015.

[31] Pradeep Natarajan, Robin Young, Nathan O Stitziel, Sandosh Padmanabhan, Usman Baber, Roxana Mehran, Samantha Sartori, Valentin Fuster, Dermot F Reilly, Adam Butterworth, et al. Polygenic risk score identifies subgroup with higher burden of atherosclerosis and greater relative benefit from statin therapy in the primary prevention setting. Circulation, 135(22):2091–2101, 2017.

[32] Amy Damask, P Gabriel Steg, Gregory G Schwartz, Michael Szarek, Emil Hagström, Lina Badimon, M John Chapman, Catherine Boileau, Sotirios Tsimikas, Henry N Ginsberg, et al. Patients with high genome-wide polygenic risk scores for coronary artery disease may receive greater clinical benefit from alirocumab treatment in the ODYSSEY OUTCOMES trial. Circulation, 141(8):624–636, 2020.

[33] Nicholas A Marston, Frederick K Kamanu, Francesco Nordio, Yared Gurmu, Carolina Roselli, Peter S Sever, Terje R Pedersen, Anthony C Keech, Huei Wang, Armando Lira Pineda, et al. Predicting benefit from evolocumab therapy in patients with atherosclerotic disease using a genetic risk score: results from the fourier trial. Circulation, 141(8):616–623, 2020.

[34] Barry J Materson, Domenic J Reda, and David W Williams. Comparison of effects of antihypertensive drugs on heart rate: changes from baseline by baseline group and over time. American journal of hypertension, 11(5):597–601, 1998.

[35] Joelle Mbatchou, Leland Barnard, Joshua Backman, Anthony Marcketta, Jack A Kosmicki, Andrey Ziyatdinov, Christian Benner, Colm O’Dushlaine, Mathew Barber, Boris Boutkov, et al. Computationally efficient whole-genome regression for quantitative and binary traits. Nature genetics, 53(7):1097–1103, 2021.

[36] Christopher C Chang, Carson C Chow, Laurent CAM Tellier, Shashaank Vattikuti, Shaun M Purcell, and James J Lee. Second-generation plink: rising to the challenge of larger and richer datasets. Gigascience, 4(1):s13742–015, 2015.

[37] Hannah V Meyer. plinkQC: genotype quality control in genetic association studies, 2022. meyerlab-cshl/plinkQC, 10.5281/zenodo.3934294.

[38] UK10K, et al. The UK10K project identifies rare variants in health and disease. Nature, 526(7571):82, 2015.

[39] Olga Krasheninina, Yih-Chii Hwang, Xiaodong Bai, Aleksandra Zalcman, Evan Maxwell, Jeffrey G Reid, and William J Salerno Jr. Open-source mapping and variant calling for large-scale ngs data from original base-quality scores. bioRxiv, pages 2020–12, 2020.

[40] Kevin J Galinsky, Gaurav Bhatia, Po-Ru Loh, Stoyan Georgiev, Sayan Mukherjee, Nick J Patterson, and Alkes L Price. Fast principal-component analysis reveals convergent evolution of adh1b in europe and east asia. The American Journal of Human Genetics, 98(3):456–472, 2016.

[41] PGS Catalog Team. PGS Catalog Calculator v2.0, 2023. https://github.com/PGScatalog/pgsc_calc.

[42] Samuel A Lambert, Laurent Gil, Simon Jupp, Scott C Ritchie, Yu Xu, Annalisa Buniello, Aoife McMahon, Gad Abraham, Michael Chapman, Helen Parkinson, et al. The polygenic score catalog as an open database for reproducibility and systematic evaluation. Nature Genetics, 53(4):420– 425, 2021.

[43] Florian Privé, Hugues Aschard, Shai Carmi, Lasse Folkersen, Clive Hoggart, Paul F O’Reilly, and Bjarni J Vilhjálmsson. Portability of 245 polygenic scores when derived from the uk biobank and applied to 9 ancestry groups from the same cohort. The American Journal of Human Genetics, 109(1):12–23, 2022.

[44] Andrew D Grotzinger, Mijke Rhemtulla, Ronald de Vlaming, Stuart J Ritchie, Travis T Mallard, W David Hill, Hill F Ip, Riccardo E Marioni, Andrew M McIntosh, Ian J Deary, et al. Genomic structural equation modelling provides insights into the multivariate genetic architecture of complex traits. Nature human behaviour, 3(5):513–525, 2019.

[45] Sarah E Graham, Shoa L Clarke, Kuan-Han H Wu, Stavroula Kanoni, Greg JM Zajac, Shweta Ramdas, Ida Surakka, Ioanna Ntalla, Sailaja Vedantam, Thomas W Winkler, et al. The power of genetic diversity in genome-wide association studies of lipids. Nature, 600(7890):675–679, 2021.

[46] Evangelos Evangelou, Helen R Warren, David Mosen-Ansorena, Borbala Mifsud, Raha Pazoki, He Gao, Georgios Ntritsos, Niki Dimou, Claudia P Cabrera, Ibrahim Karaman, et al. Genetic analysis of over 1 million people identifies 535 new loci associated with blood pressure traits. Nature genetics, 50(10):1412–1425, 2018.

