## Supplementary Material for "Leveraging large-scale biobank EHRs to enhance pharmacogenetics of cardiometabolic disease medications"

### 1 Supplementary Notes

#### Supplementary Note 1.

According to Figure 3a, biomarker levels $Y$ at time $t$ can be modelled as follows:

$$Y_t = \beta_0 \cdot G_0 + \beta_E \cdot E_t + \gamma_E \cdot G_E \cdot E_t + \beta_D \cdot D_t + \gamma_D \cdot G_D \cdot D_t + \epsilon_t \tag{1}$$

where $\beta_0$ is the baseline genetic effect, $G$ the genetics, $\beta_E$ the environmental effect, $E$ the environment, $\gamma_E$ the gene-environment interaction effect, $D$ the indicator of drug use, $\beta_D$ the drug effect and $\gamma_D$ the pharmacogenetic effect.

The drug response phenotype which is the difference between post-treatment $Y_{t_1}$ and baseline $Y_{t_0}$ biomarker levels can thus be modelled as follows:

$$Y_{t_1} - Y_{t_0} = \beta_E(E_{t_1} - E_{t_0}) + \gamma_E \cdot G_E(E_{t_1} - E_{t_0}) + \beta_D(D_{t_1} - D_{t_0}) + \gamma_D \cdot G_D(D_{t_1} - D_{t_0}) + \Delta\epsilon_{01} \tag{2}$$

$$= \beta_D + \gamma_D \cdot G_D + \delta_{01} \tag{3}$$

where baseline genetics of $Y_{t_1}$ and $Y_{t_0}$ cancel each other out, $D_{t_1}$ and $D_{t_0}$ correspond by definition to 1 and 0, respectively, and $(\beta_E + \gamma_E \cdot G_E)(E_{t_1} - E_{t_0}) + \Delta\epsilon_{01}$ are regrouped under $\delta_{01}$ assuming that interactions between genetics and changing environments are negligible (see control GWAS on longitudinal change in Figures SS9-SS10). The same derivation applies to the logarithmic difference $log(Y_{t_1}) - log(Y_{t_0}) = log(Y_{t_1}/Y_{t_0})$ which can be interpreted as a relative change in biomarker levels.

Adjusting drug response phenotypes or change scores for baseline biomarker levels $Y_{t_0}$ wrongly introduces baseline genetic effects into expression 3 which results in the estimation of $\beta_0$ in addition to $\gamma_D$ which we elaborate in the following.

To simplify the calculations, let us assume that all these variables ($Y$, $G$, $E$) are scaled to have zero mean and unit variance. When we regress $Y_{t_1}$ onto $Y_{t_0}$ the regression estimate $\widehat{\alpha}$ will be

$$\begin{aligned}
E[\widehat{\alpha}] = E[Y_{t_1} \cdot Y_{t_0}] &= E[(\beta_0 \cdot G_0 + \beta_E \cdot E_{t_1} + \gamma_E \cdot G_E \cdot E_{t_1} + \beta_D \cdot D_{t_1} + \gamma_D \cdot G_D \cdot D_{t_1} + \epsilon_{t_1}) \\
&\times (\beta_0 \cdot G_0 + \beta_E \cdot E_{t_0} + \gamma_E \cdot G_E \cdot E_{t_0} + \beta_D \cdot D_{t_0} + \gamma_D \cdot G_D \cdot D_{t_0} + \epsilon_{t_0})] \\
&= \beta_0^2 + \beta_E^2 \cdot corr(E_{t_1}, E_{t_0}) + \gamma_E^2 \cdot var(G_E) \cdot corr(E_{t_1}, E_{t_0}) \\
&= \beta_0^2 + (\beta_E^2 + \gamma_E^2) \cdot corr(E_{t_1}, E_{t_0})
\end{aligned}$$

Thus the residual from such a regression will be

$$R_{0,1} = Y_{t_1} - \widehat{\alpha} \cdot Y_{t_0} \tag{4}$$

Note that $\widehat{\alpha}$ only changes by a constant 1 if we regress the biomarker difference $Y_{t_1} - Y_{t_0}$ onto $Y_{t_0}$ instead of $Y_{t_1}$ onto $Y_{t_0}$, making these two approaches equivalent. This can be shown as follows:

$$Y_{t_1} - Y_{t_0} = \alpha' \cdot Y_{t_0} + \epsilon \tag{5}$$

Rearranging Equation 5 results in:

$$Y_{t_1} = (\alpha' + 1) \cdot Y_{t_0} + \epsilon = \alpha \cdot Y_{t_0} + \epsilon \tag{6}$$

When running a GWAS on this residual response phenotype its correlation with $G_0$ will be

$$corr(R_{0,1}, G_0) = E[(Y_{t_1} - \widehat{\alpha} \cdot Y_{t_0}) \cdot G_0] = \beta_0^2 \cdot (1 - \widehat{\alpha}) = \beta_0^2 \cdot (1 - \beta_0^2 - (\beta_E^2 + \gamma_E^2) \cdot corr(E_{t_1}, E_{t_0})) \tag{7}$$

This means that when regressing the post-treatment effect on the pre-treatment effect and running a GWAS on the residuals, we expect to see a strong (spurious) genetic correlation with the genetic basis of the (time-invariant) baseline effect. Note that this correlation between the residual and the baseline genetics is identical whether we used drug-naive or pre- vs post-treated individuals. This observation further confirms that genetic “discoveries” based on residual response definitions are likely to be non-specific to the treatment.

If we examine the correlation between these residuals and the underlying drug response genetics we have

$$corr(R_{0,1}, G_D) = E[(Y_{t_1} - \widehat{\alpha} \cdot Y_{t_0}) \cdot G_0] = \gamma_D \cdot D_{t_1} - \widehat{\alpha} \cdot \gamma_D \cdot D_{t_0} \tag{8}$$

Thus, this correlation in drug-naive samples (where $D_{t_0} = D_{t_1} = 0$) is zero, but in post- vs pre-treated samples (where $D_{t_0} = 0$ and $D_{t_1} = 1$) is $\gamma_D$.

It is clear that if we simply use the post-treatment vs baseline difference, i.e. $Y_{t_1} - Y_{t_0}$, its correlation with $G_0$ is zero and its correlation with $G_D$ is $\gamma_D$. Therefore, it is strongly recommended to use the simple post-treatment - baseline biomarker difference to elucidate the pure treatment-specific genetic effects.

#### Supplementary Note 2.

HbA1c values were either DCCT (Diabetes Control and Complications Trial) aligned (codes: 42W4. and XaERp; percentage unit) or IFCC (International Federation of Clinical Chemistry and Laboratory Medicine) aligned (42W5. and XaPbt; mmol/mol unit). For consistency, we used mmol/mol units and converted DCCT units using the NGSP/IFCC equation recommended by the National Glycohemoglobin

Standardization Program (NGSP) network (`https://ngsp.org/ifcc.asp`): NGSP = [0.09148 · IFCC] + 2.152.

#### Supplementary Note 3.

Medication codes can correspond to multiple active ingredients taken in combination, among which the primary medication of interest. Since we cannot disentangle the effect of the primary medication compared to a second ingredient taken in combination, we filter out individuals with prescriptions corresponding to combination therapies during the study period. For statins, we eliminate combination therapies with ezetimibe and fenofibrate, for metformin, combination therapies with sitagliptin, linagliptin, saxagliptin, alogliptin, dapagliflozin, canagliflozin, empagliflozin, rosiglitazone, pioglitazone, vildagliptin and for beta blockers, combination therapies with diuretics and aspirin.

Note that this step is specific to drugs with a combined formulation and is different from the QC step where individuals taking a drug from the same medication class, but with a separate prescription code, are filtered out.

### 2 Supplementary Figures

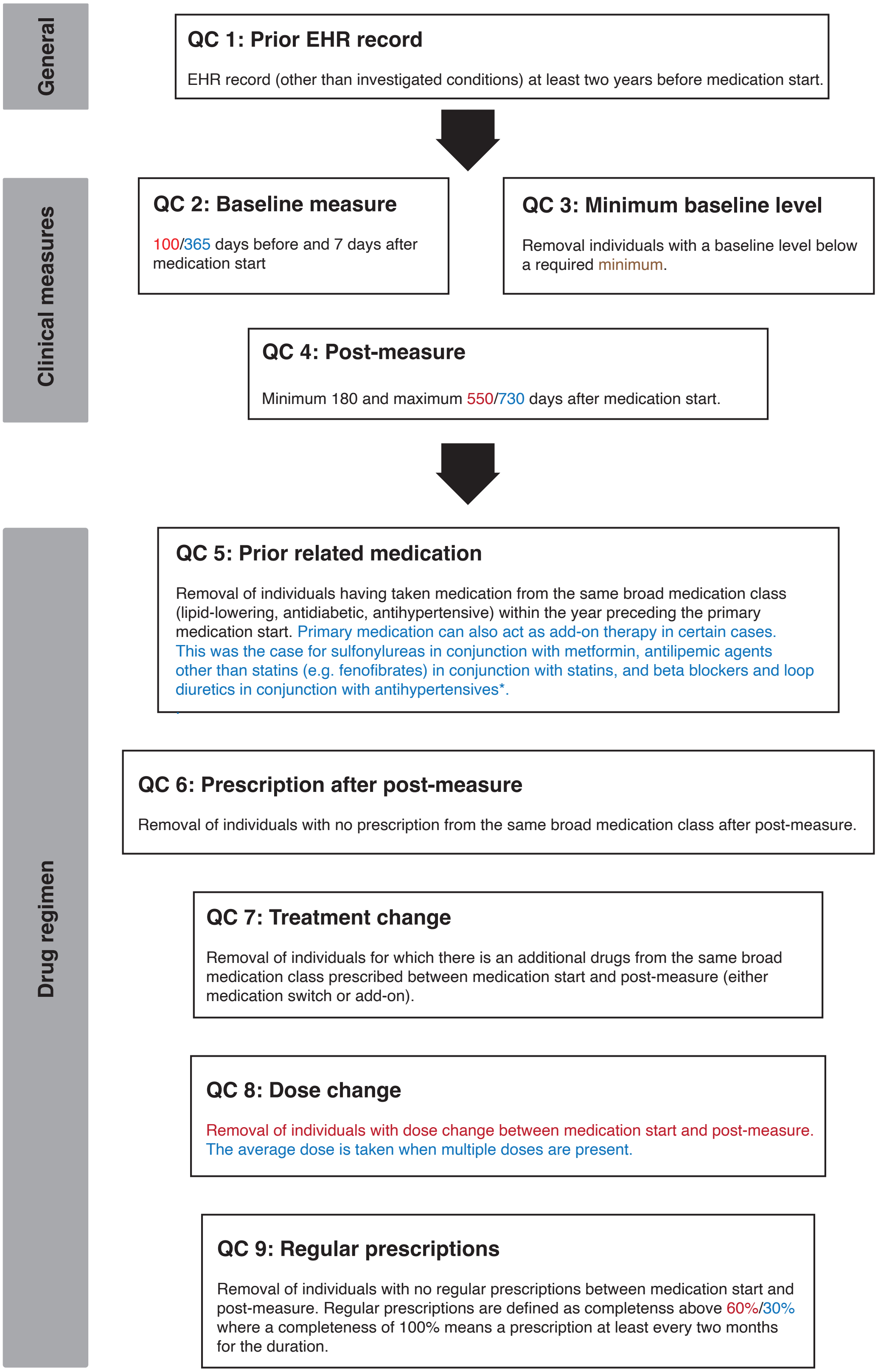

Figure S1: **Flow diagram of quality control steps.** After selecting individuals taking the primary medication of interest, individuals with missing biomarker measures, medication therapy changes before post-treatment measures, irregular prescriptions, or not enrolled in the health care system before the medication start were removed. Stringent filtering criteria are written in red and lenient ones in blue. Medication/phenotype-specific criteria are written in brown.

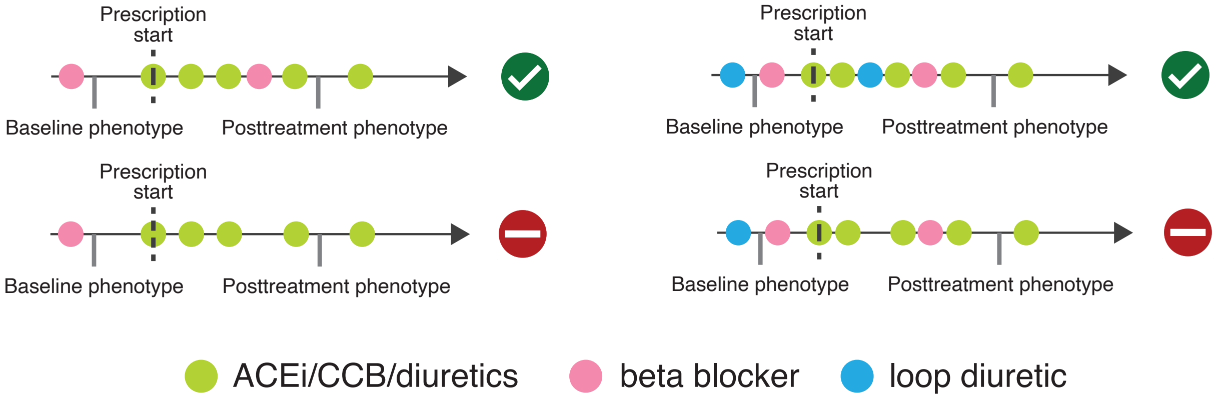

Figure S2: **Add-on therapy definition.** For antihypertensives, primary medication (ACEi, CCB and thiazide diuretics) could also act as add-on therapy to beta blockers and loop diuretics. However, medication prescribed before the primary medication start was also required to be prescribed afterwards (at least until the post-treatment measure). If the start of a beta blocker or loop diuretics medication was after the prescription start of the primary medication, this would count as "treatment change" and the individual would be removed.

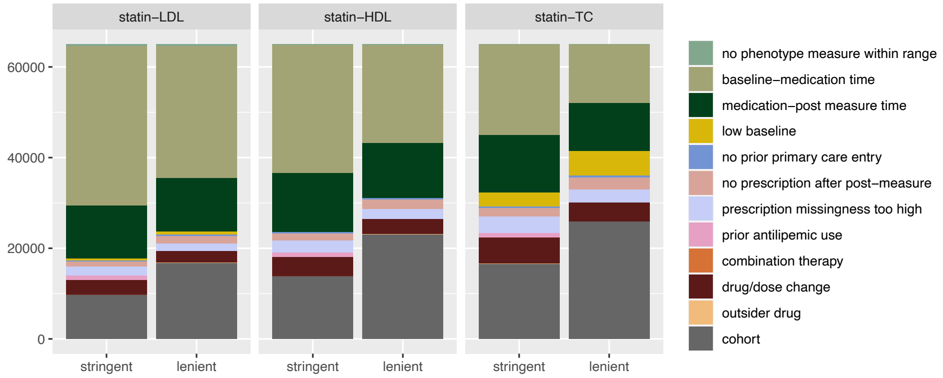
statin–LDL
statin–HDL
statin–TC
60000
40000
20000
0
stringent
lenient
no phenotype measure within range
baseline–medication time
medication–post measure time
low baseline
no prior primary care entry
no prescription after post–measure
prescription missingness too high
prior antilipemic use
combination therapy
drug/dose change
outsider drug
cohort

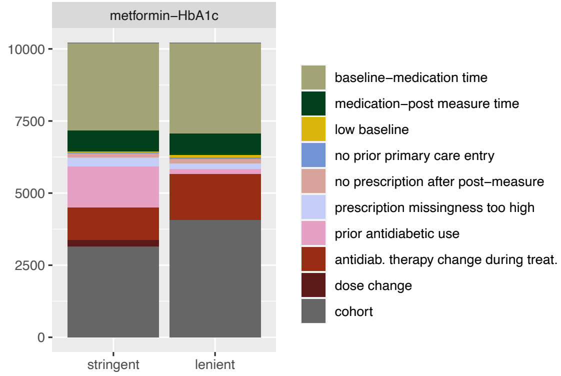
metformin–HbA1c
10000
7500
5000
2500
0
stringent
lenient
baseline–medication time
medication–post measure time
low baseline
no prior primary care entry
no prescription after post–measure
prescription missingness too high
prior antidiabetic use
antidiab. therapy change during treat.
dose change
cohort

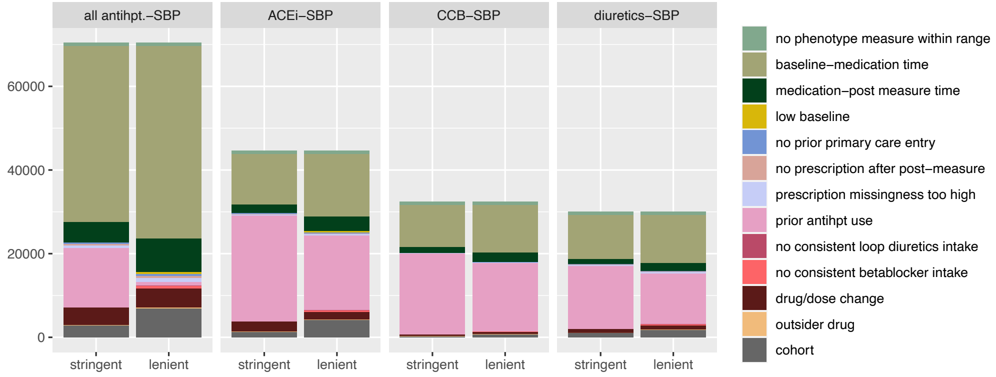
all antihpt.–SBP
ACEi–SBP
CCB–SBP
diuretics–SBP
60000
40000
20000
0
stringent
lenient
no phenotype measure within range
baseline–medication time
medication–post measure time
low baseline
no prior primary care entry
no prescription after post–measure
prescription missingness too high
prior antihpt use
no consistent loop diuretics intake
no consistent betablocker intake
drug/dose change
outsider drug
cohort

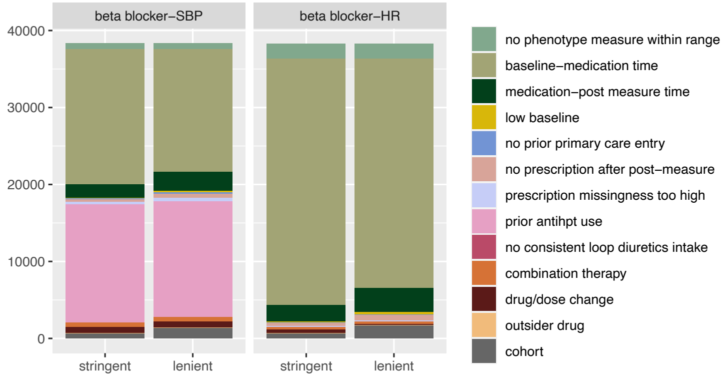
beta blocker–SBP
beta blocker–HR
40000
30000
20000
10000
0
stringent
lenient
no phenotype measure within range
baseline–medication time
medication–post measure time
low baseline
no prior primary care entry
no prescription after post–measure
prescription missingness too high
prior antihpt use
no consistent loop diuretics intake
combination therapy
drug/dose change
outsider drug
cohort

Figure S3: **Number of individuals in each UK Biobank drug response cohort and reasons for removal (stacked barplot).** The height of the bar represents the number of individuals having at least one prescription of the investigated drug. The bottom grey bar represents the number of individuals after QC steps. Note that some filtering reasons are not mutually exclusive. For instance, baseline-medication time filtering was done after checking for prior related medication. Therefore, for metformin-HbA1c in the lenient scenario, it seems that more individuals were filtered out because of baseline-medication time than in the stringent scenario. However, given that individuals with previous sulfonylureas use were excluded in the stringent, but included in the lenient filtering setting, there is a larger pool of individuals in the lenient scenario for whom baseline measures are potentially missing. The same reasoning holds for antihypertensives where individuals with prior beta blocker and loop diuretics prescriptions were included in the lenient filtering scenario (see Figure S2).

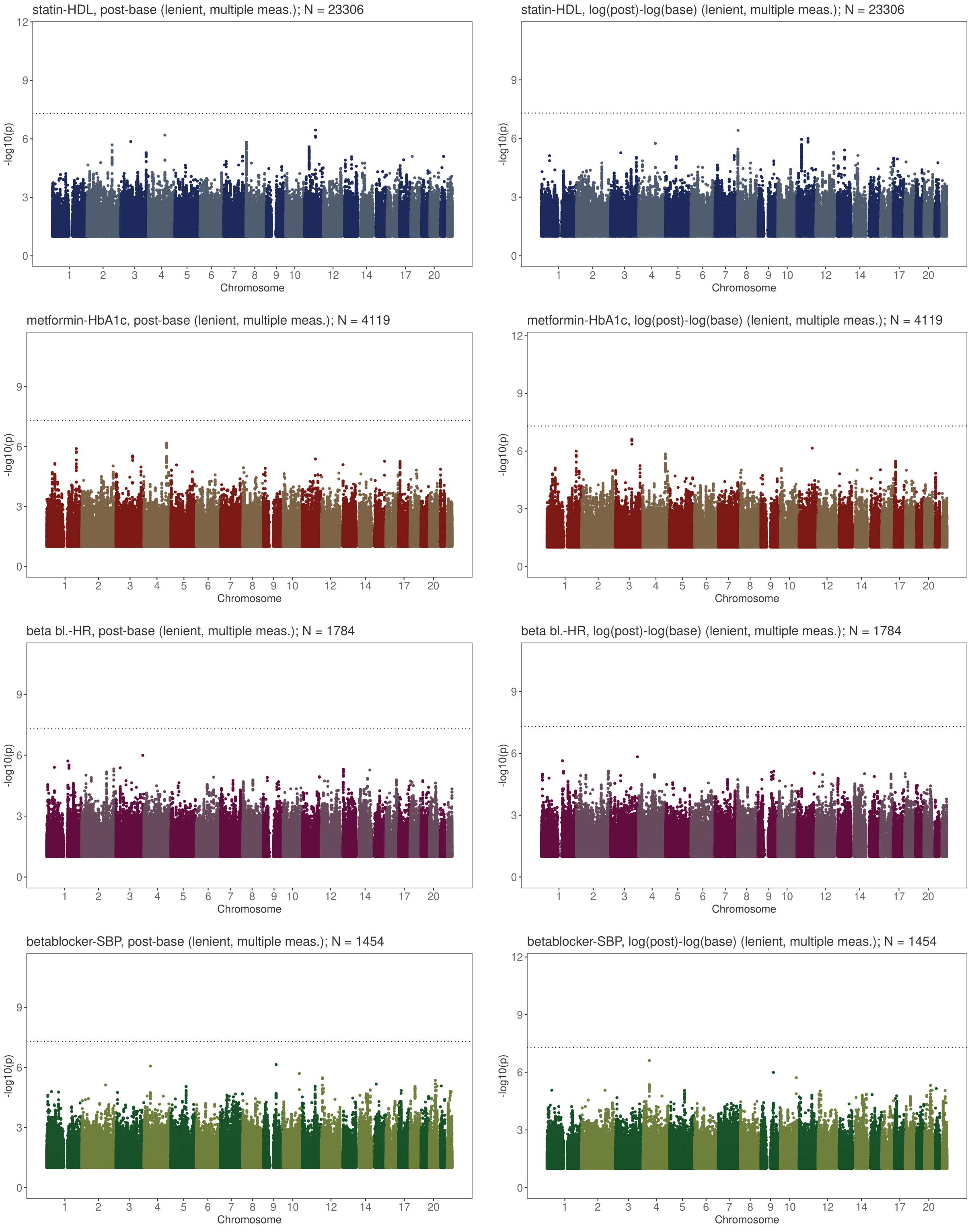

Figure S4: **HbA1c response to metformin, SBP response to beta blocker and HR response to beta blocker GWAS results.** Plots on the left show GWAS results for the absolute biomarker (post-baseline level) and plots on the right the results for the logarithmic relative (log(post)-log(base)) difference. GWAS results correspond to the lenient filtering scenarios with average baseline and post-treatment values over multiple measures if available. Genome-wide significant loci are annotated with the closest gene. The horizontal line denotes genome-wide significance (p-value $<$ 5e-8).

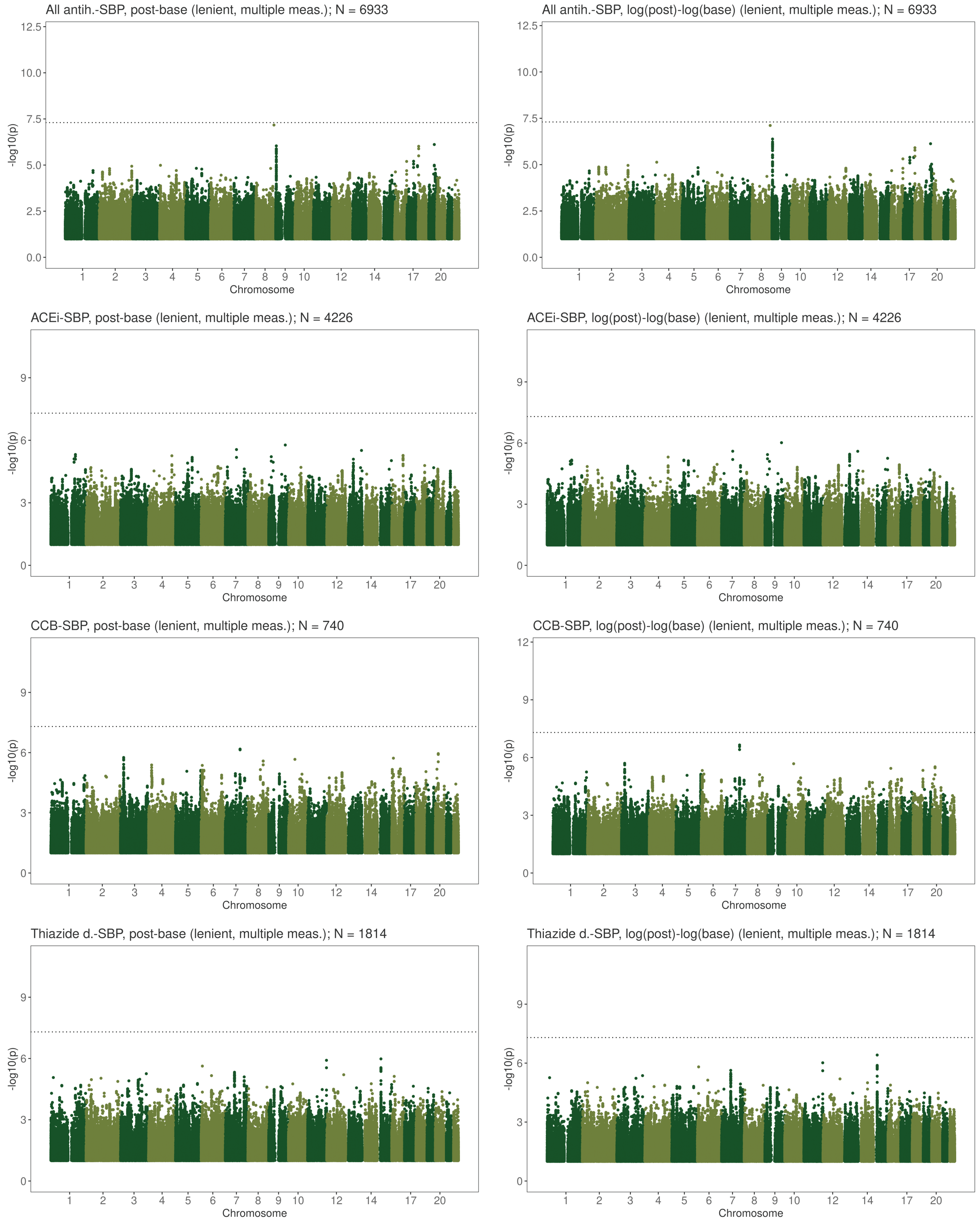

Figure S5: **SBP response to first-line antihypertensives GWAS results.** Plots on the left show GWAS results for the absolute biomarker (post-baseline level) and plots on the right the results for the logarithmic relative (log(post)-log(base)) difference. GWAS results correspond to the lenient filtering scenarios with average baseline and post-treatment values over multiple measures if available. Genome-wide significant loci are annotated with the closest gene. The horizontal line denotes genome-wide significance (p-value $<$ 5e-8).

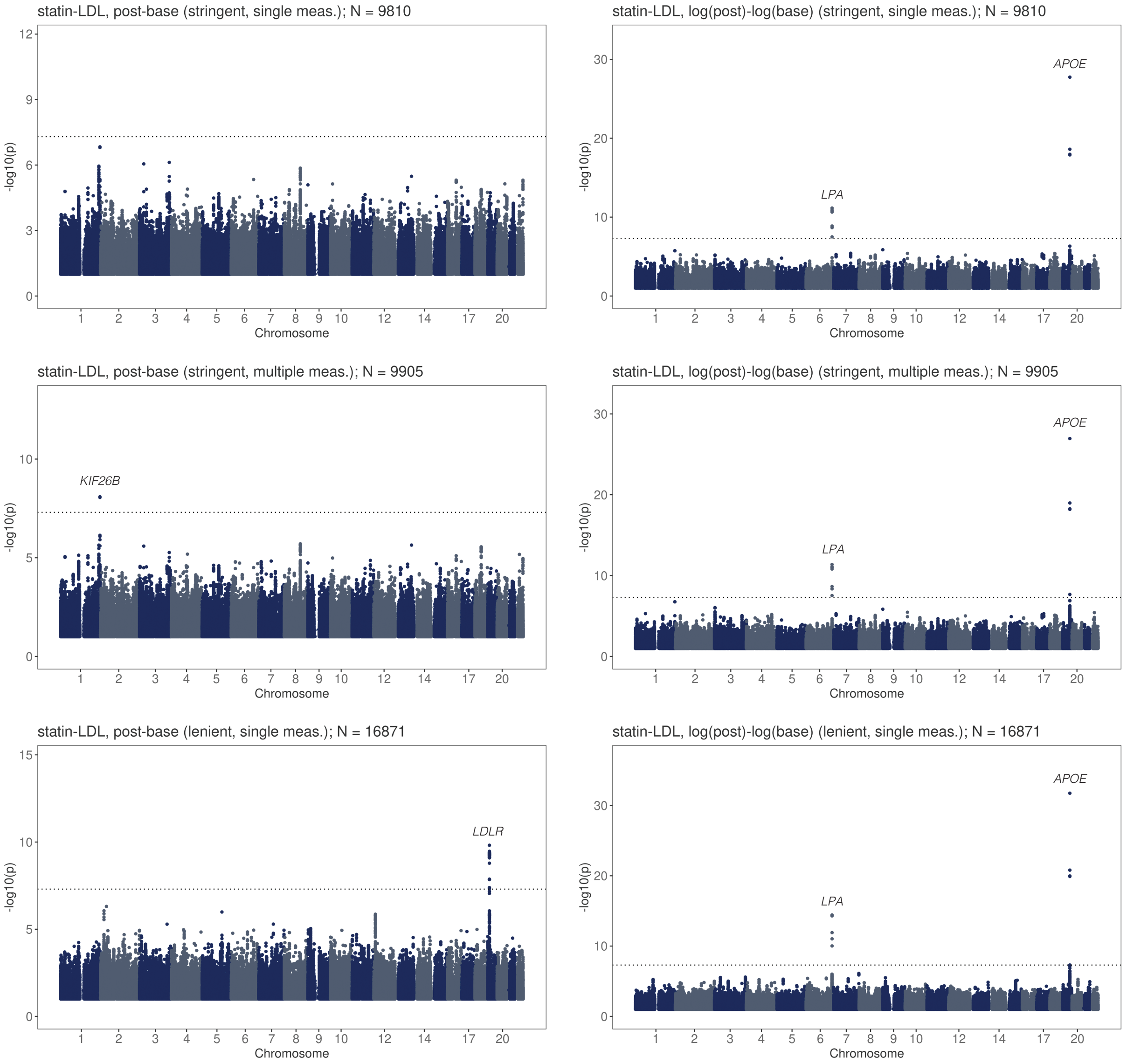

Figure S6: **LDL-C response to statins GWAS results in the different filtering scenarios.** Plots on the left show GWAS results for the absolute biomarker (post-baseline level) and plots on the right the results for the logarithmic relative (log(post)-log(base)) difference. For stringent and lenient filtering scenarios, single baseline and post-treatment measures and average values over multiple measures, if available, were tested. Results for lenient filtering and multiple measures are shown in Figure 3. Genome-wide significant loci are annotated with the closest gene. The horizontal line denotes genome-wide significance (p-value $<$ 5e-8).

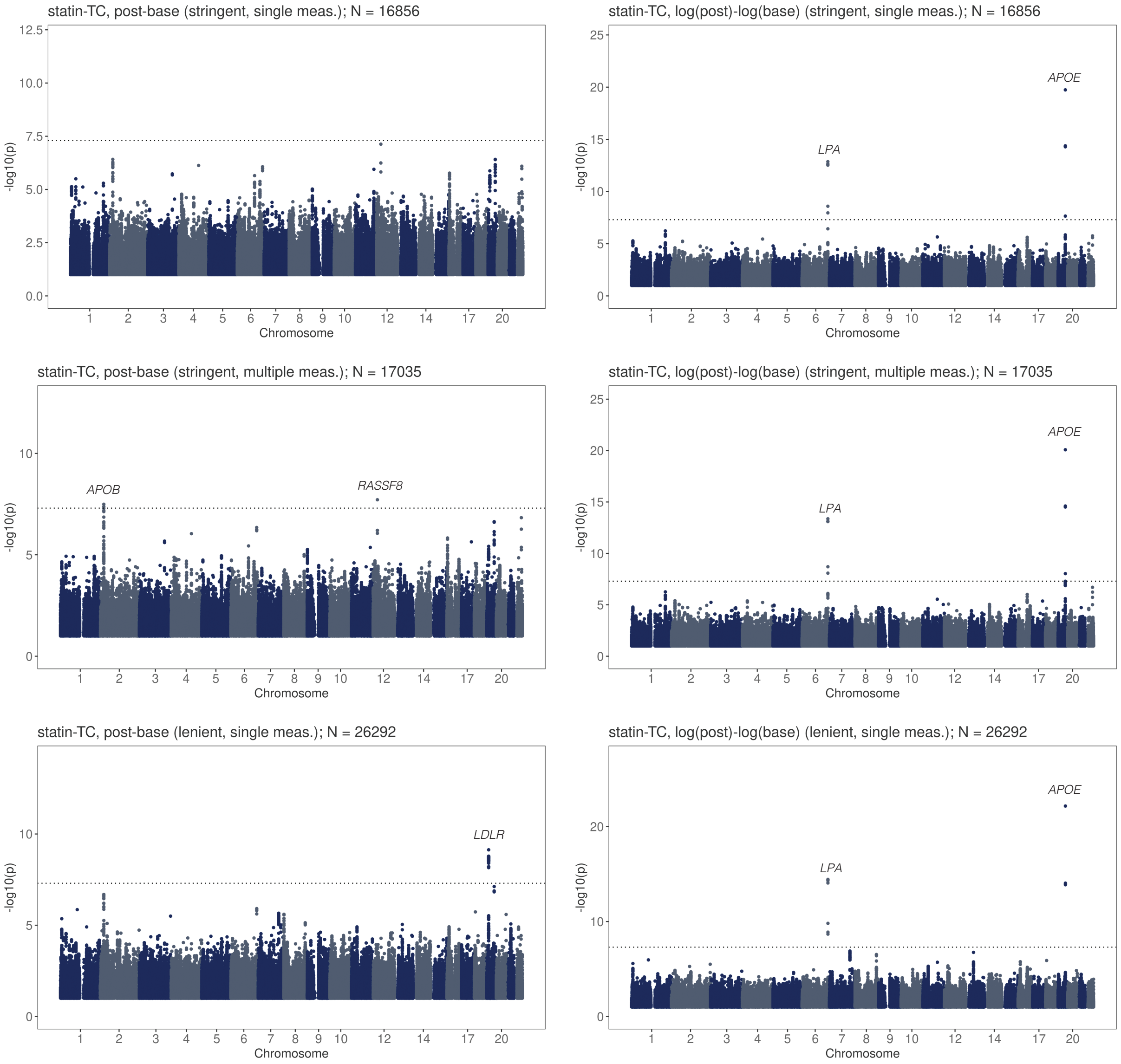

Figure S7: **Total cholesterol (TC) response to statins GWAS results in the different filtering scenarios.** Plots on the left show GWAS results for the absolute biomarker (post-baseline level) and plots on the right the results for the logarithmic relative (log(post)-log(base)) difference. For stringent and lenient filtering scenarios, single baseline and post-treatment measures and average values over multiple measures, if available, were tested. Results for lenient filtering and multiple measures are shown in Figure 3. Genome-wide significant loci are annotated with the closest gene. The horizontal line denotes genome-wide significance (p-value $<$ 5e-8).

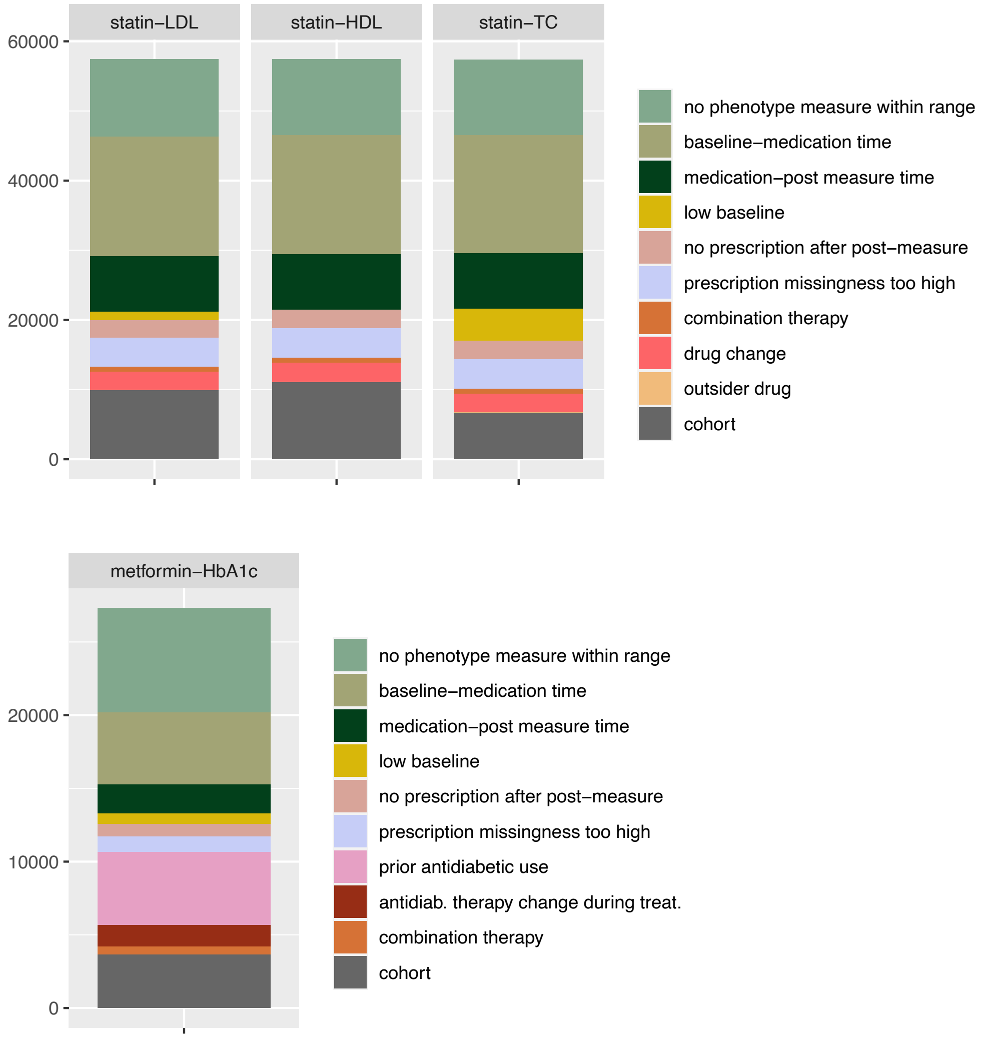

Figure S8: **Number of individuals in each All of Us drug response cohort and reasons for removal (stacked barplot).** The height of the bar represents the number of individuals having at least one prescription of the investigated drug. The bottom grey bar represents the number of individuals after QC steps. Note some filtering reasons are not mutually exclusive.

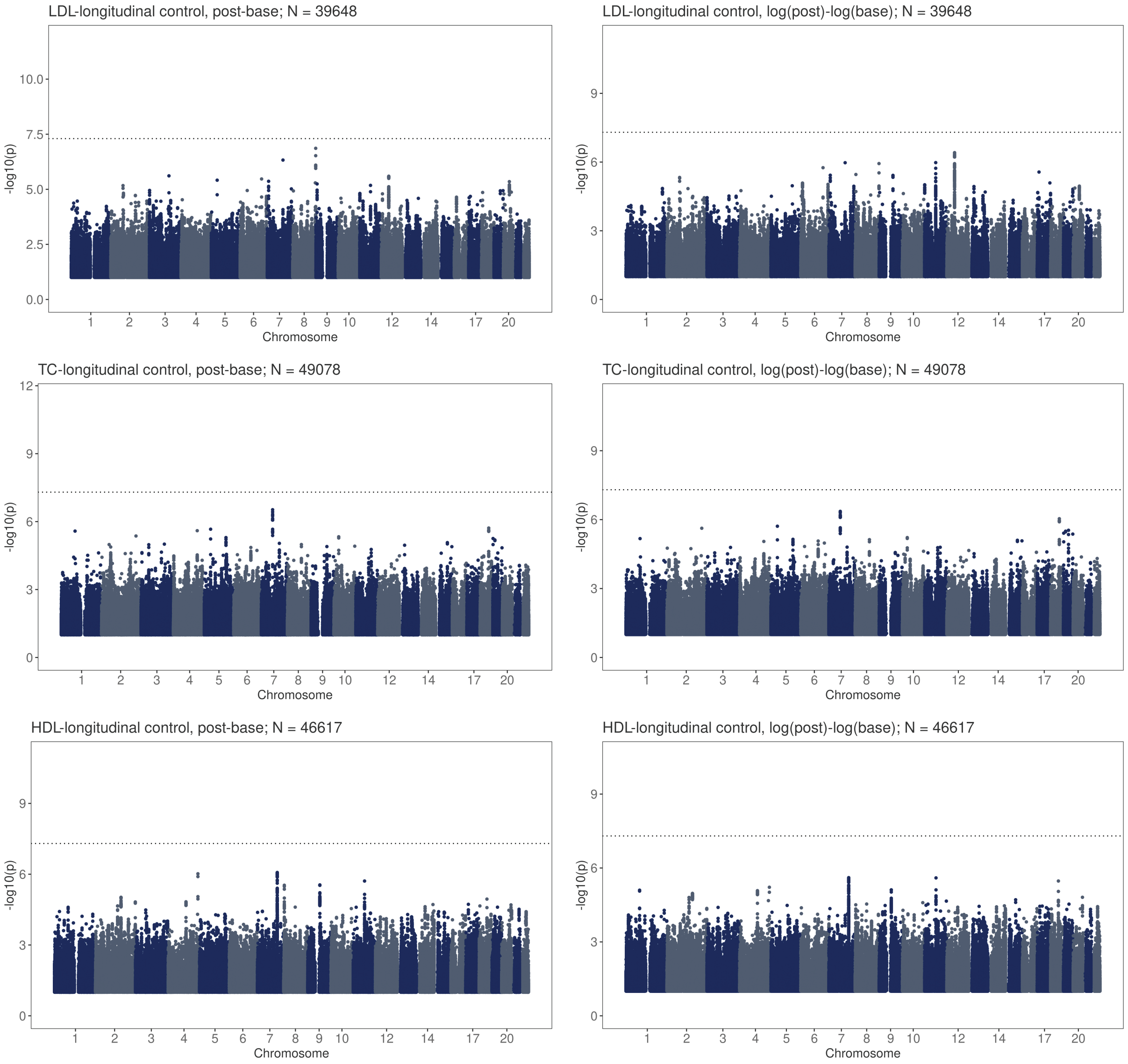

Figure S9: **Longitudinal biomarker change GWAS in medication-naive individuals for LDL, TC and HDL.** Plots on the left show GWAS results for the absolute biomarker (post-baseline level) and plots on the right the results for the logarithmic relative (log(post)-log(base)) difference. Genome-wide significant loci are annotated with the closest gene. The horizontal line denotes genome-wide significance (p-value < 5e-8).

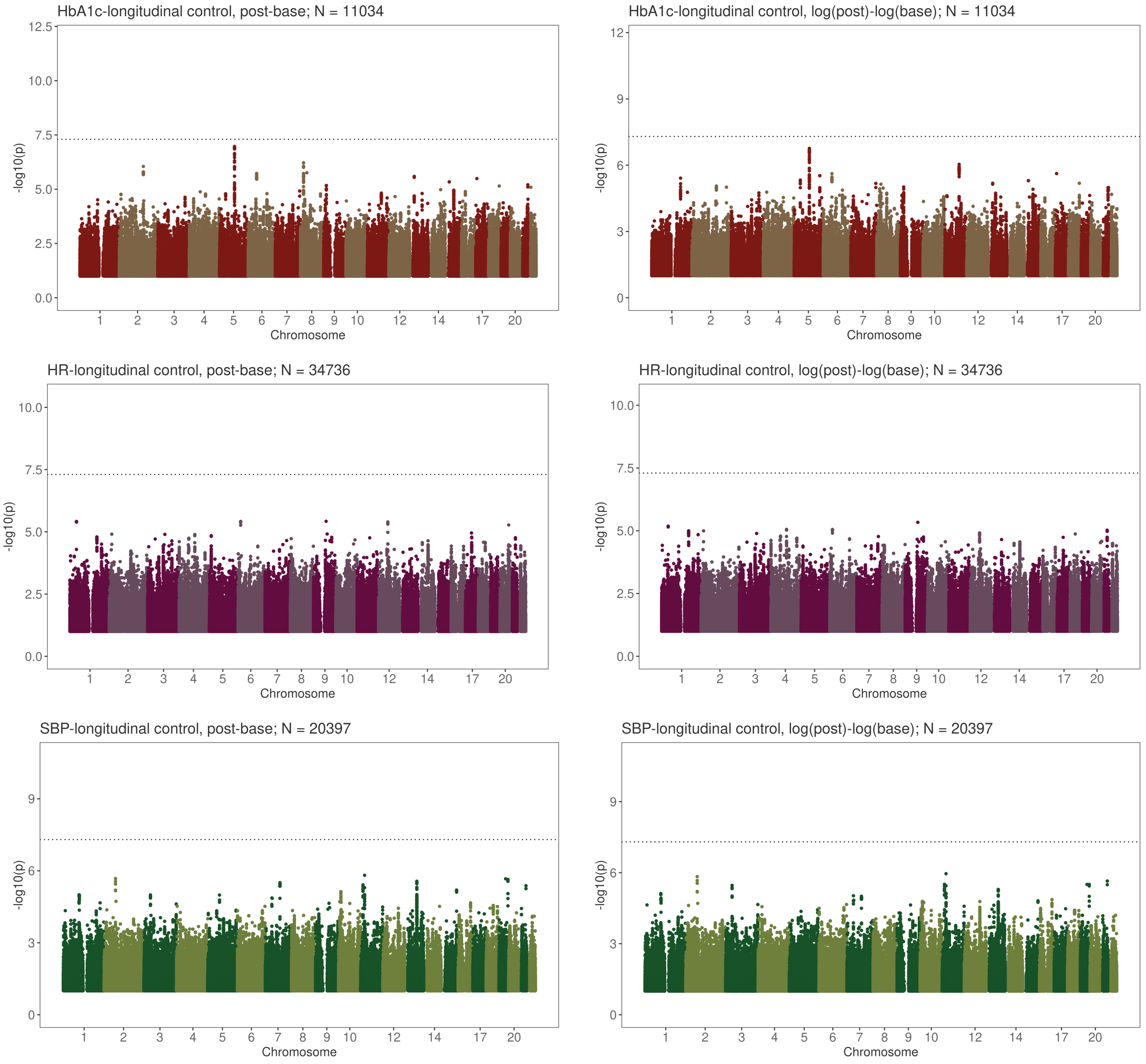

Figure S10: **Longitudinal biomarker change GWAS in medication-naive individuals for HbA1c, HR and SBP.** Plots on the left show GWAS results for the absolute biomarker (post-baseline level) and plots on the right the results for the logarithmic relative (log(post)-log(base)) difference. Genome-wide significant loci are annotated with the closest gene. The horizontal line denotes genome-wide significance (p-value $<$ 5e-8).

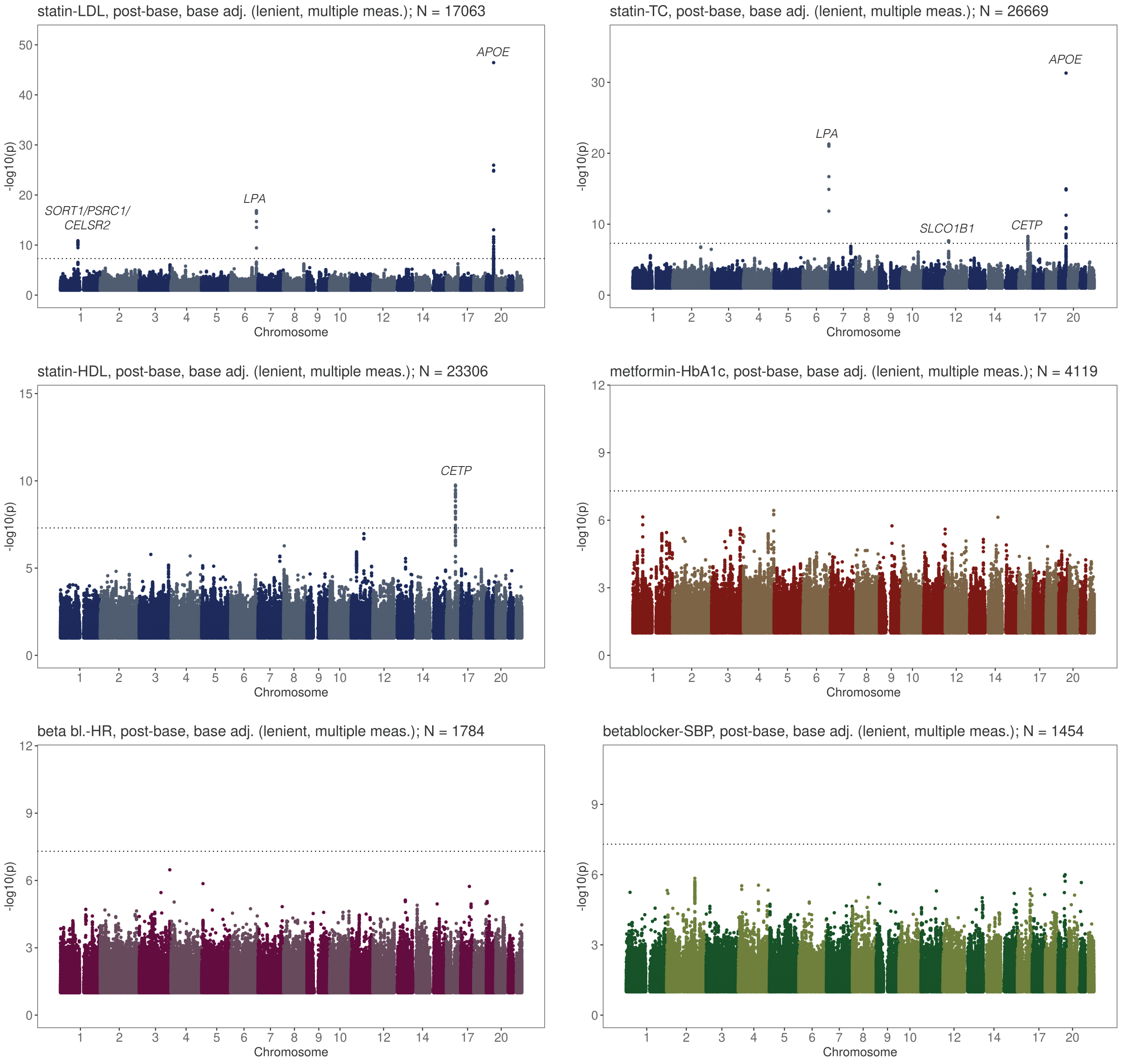

Figure S11: **GWAS results for baseline adjusted drug response phenotypes (statin-LDL, statin-TC, statin-HDL, metformin-HbA1c, beta blocker-HR, beta blocker-SBP).** GWAS results correspond to the lenient filtering scenarios with average baseline and post-treatment values over multiple measures if available. Genome-wide significant loci are annotated with the closest gene. The horizontal line denotes genome-wide significance (p-value $<$ 5e-8).

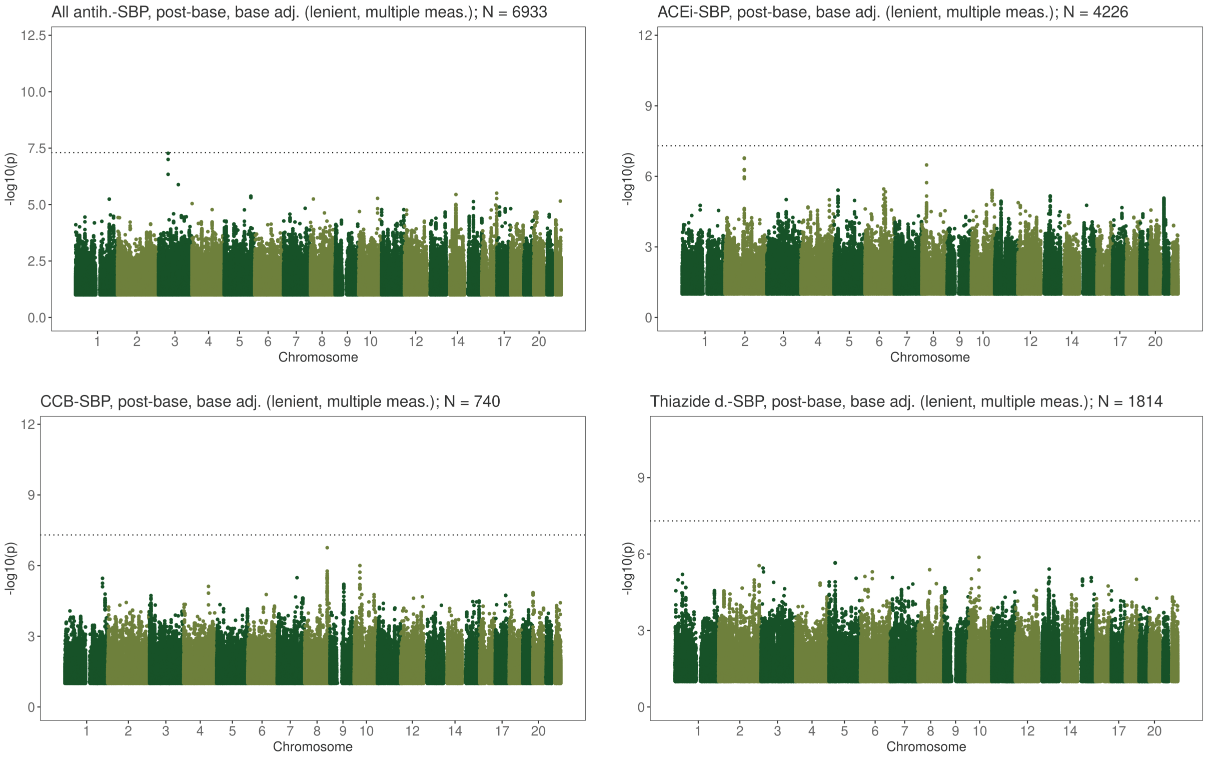

Figure S12: **GWAS results for baseline adjusted drug response phenotypes (SBP response to first-line antihypertensives).** GWAS results correspond to the lenient filtering scenarios with average baseline and post-treatment values over multiple measures if available. Genome-wide significant loci are annotated with the closest gene. The horizontal line denotes genome-wide significance (p-value $<$ 5e-8).

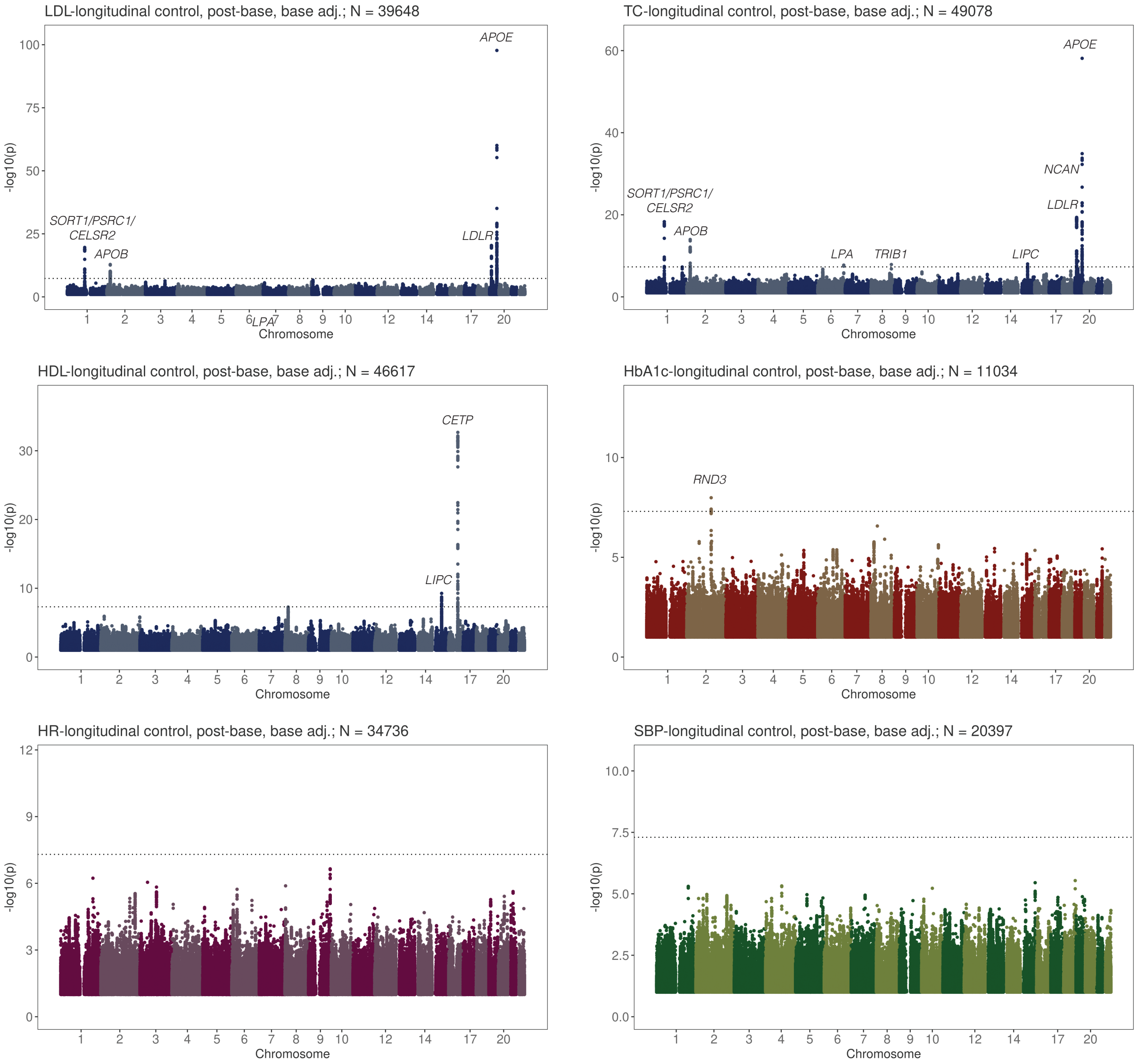

Figure S13: **Baseline adjusted longitudinal biomarker change GWAS in medication-naive individuals.** Genome-wide significant loci are annotated with the closest gene. The horizontal line denotes genome-wide significance (p-value $<$ 5e-8).

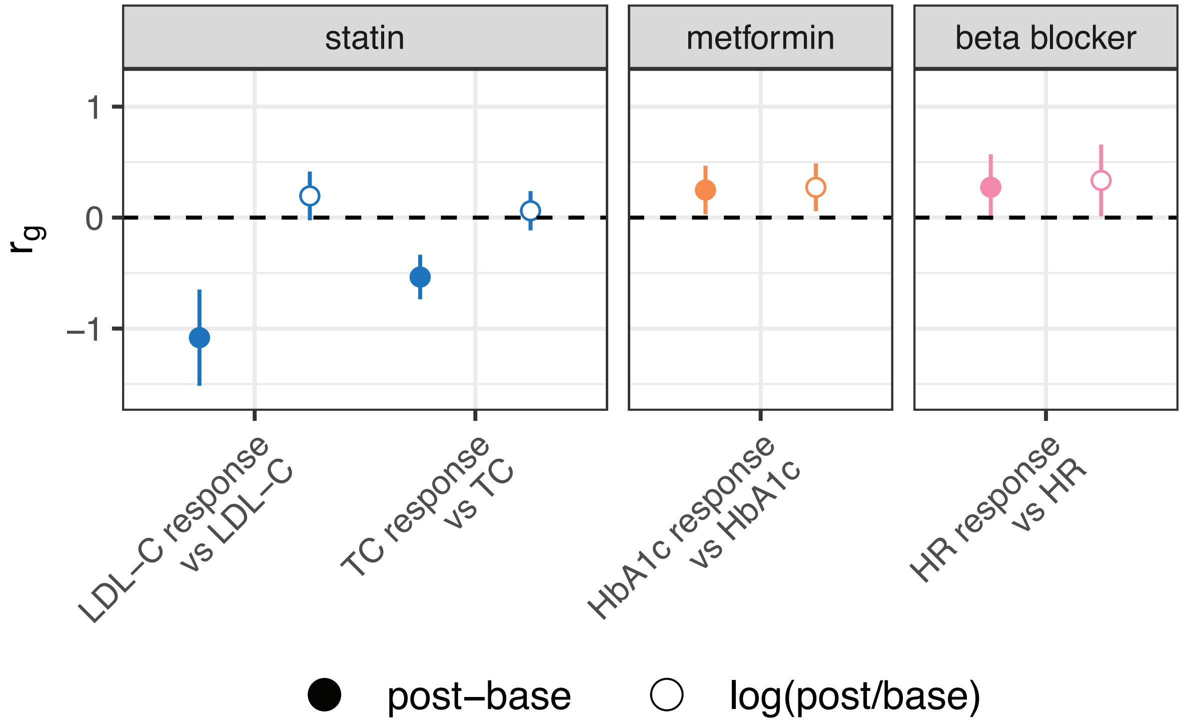

Figure S14: **Genetic correlations ($r_g$) between drug response and underlying baseline traits.** Error bars correspond to the standard error. Drug response phenotypes with nominally significant genetic correlations for at least one of the post-base and log(post/base) definitions are shown. Numerical values are in Table S13.
